# Coherence-Sense experiencing of Ukrainian Forced Migrants in The Conditions of a Full-Scale Russian Military Invasion: Behavioral Patterns, Mental Health, Believes and Everyday Challenges

**DOI:** 10.1101/2024.06.29.24309715

**Authors:** Vitalii Lunov

## Abstract

Considering the Russia-Ukraine war and its profound impact on displaced populations, this study investigates the psychological, behavioral, and cultural characteristics of Ukrainian forced migrants, emphasizing gender-based differences. Using a diverse set of assessment tools, the research explores how male and female migrants cope with the complex challenges of displacement, focusing on their sense of coherence, mental health, physical well-being, learning behaviors, work attitudes, self-organization, cultural values, and religiosity. *Materials and Methods*. The study sampled 742 Ukrainian migrants, including 253 males and 489 females, from various European countries. Assessments included the Sense of Coherence Scale (SOC), Screening-Questionnaire of Negative and Positive Symptoms, Health Functionality Questionnaire, Learning Behavior Survey, Work Attitude Test, Self-Organization Questionnaire, Cultural Value Differential, and Religiosity Structure Test. Statistical analyses were performed using SPSS-28, incorporating factor analysis to identify key trends and differences. *Results.* Key findings reveal higher SOC scores among male migrants, indicating better life manageability and meaningfulness. Female migrants report higher levels of anxiety, depressive states, and health issues, reflecting greater emotional and physical burdens. Differences in learning behaviors show females face more organizational challenges but exhibit higher learning motivation. Male migrants demonstrate greater work engagement and regularity in self-organization, while females emphasize respect for authority and traditional values. Religiosity patterns indicate females derive emotional support from faith, whereas males use religiosity for identity and community bonding. *Conclusions.* The study underscores the necessity for gender-sensitive interventions to support Ukrainian forced migrants. Tailored approaches addressing the distinct psychological, behavioral, and cultural needs of male and female migrants are crucial for enhancing their well-being and integration into new communities during times of crisis.

## Introduction

Migration, a complex socio-political and psychological phenomenon, has historically catalyzed significant shifts in the global narrative. Often, individuals migrate to seek better opportunities or escape adverse conditions, but forced migrations, especially those stemming from full-scale military invasions, inject an added layer of intricacy into this already multifaceted issue. In the context of the Russian military invasion, countless Ukrainians found themselves thrust into this maelstrom of displacement, having to grapple not just with the loss of their homes, but with the intricate psychological, behavioral, and neuro-clinical ramifications of such a drastic life change.

The term "Coherence-Sense" encapsulates the essence of one’s ability to perceive life as comprehensible, manageable, and meaningful. It is a crucial construct when exploring the experiences of migrants, more so for those displaced by conflict, as it touches upon their resilience, their coping mechanisms, and the very fabric of their existential experiences in unfamiliar terrains. Integrating this with the social-behavioral challenges and the clinical-neuro-psychological patterns they manifest provides a holistic lens through which to study these migrants.

Gender, as highlighted in earlier discussions, plays a pivotal role in these dynamics, with male and female migrants often displaying different coping strategies and outcomes. Be it in the realm of behavioral symptoms, health-related quality of life, attitudes towards learning, work, and religiosity, or their self-organization amidst chaos, these gender-differentiated patterns provide a nuanced understanding of the broader migratory experience.

## Objective

The primary aim of this article is to delve deep into the multi-layered experiences of these migrants. Through a comprehensive exploration of their coherence-sense, the study seeks to unveil the intricate interplay of socio-behavioral patterns and clinical-neuro-psychological outcomes. By doing so, it aspires to shed light on the resilience, vulnerabilities, and adaptive strategies of Ukrainian migrants, ultimately providing insights that can inform policies, therapeutic interventions, and support systems tailored to bolster their well-being and integration into new societies.

## Methods and Data

In a study organized by the European Academy of Sciences of Ukraine "Ukrainian in the world: a socio-behavioral and clinical-psychological study of internally displaced persons and migrants", which was conducted on a sample of displaced persons from Ukraine to Austria, Hungary, Georgia, Italy, UAE, Poland, Romania, Slovakia, and the Czech Republic the following was established.

The study protocol was approved by the Research Ethics Commission of Department of general and medical psychology at Bogomolets national medical university (approval No. 1-29.03.2022).

The sample consisted of 253 males and 489 females, aged 28 to 53 years.

1) Sense of coherence scale – SOC (A. Antonovsky, 1987; 1993) ^i ii^
2) Screening-questionnaire of negative and positive symptoms (V. Lunov)
3) Questionnaire for assessing the level of health on the main functional systems (V. Voinov, L.Bugaev, S. Kulba, etc., 1999).^iii^
4) Attitude towards learning and acquiring new knowledge
5) Test "The attitude to work" (K. Maslach, M. Leiter, 1988)^iv^
6) Questionnaire for self-organization of activities (E. Mandrikova)^v^
7) Cultural value differential (G. Soldatova, S. Ryzhova, 1998)^vi^
8) Test to determine the structure of individual religiosity (Yu.Shcherbatykh, 1996).^vii^

## Main indicators of the impact of the war on forced migrants

19.2% of respondents report that their relatives and family members have experienced complete, and 19.5% have experienced partial destruction of housing and property. In 27.6% of Ukrainian migrants, people from close circle experienced disability, bodily injuries, physical injuries because of hostilities.

37.0 Ukrainian migrants have new somatic (physical) symptoms, but they do not significantly affect life and well-being. In 16.4% of respondents, the manifestation of this symptom is strong; 11.2% - feel exhausted and disorganized due to the appearance of new somatic symptoms.

In 38.4% of Ukrainian migrants, old somatic (physical) symptoms intensified, but they do not significantly affect life and well-being. At the same time, in 12.3%, the manifestation of this symptom is strong and 5.7% of the respondents feel exhausted and disorganized due to the intensification of somatic symptoms.

It should be noted that 26.0% of Ukrainian migrants developed problems related to mental health, while in 17.8% of respondents the manifestation of this symptom is strong.

Assessing the everyday difficulties faced by Ukrainian migrants, it was found that there are restless thoughts about the future.23.3% have certain signs of anxiety about the future, but they do not significantly affect life and well-being; in 47.9%, the manifestation of anxiety is strong; 12.3% feel exhausted and disorganized when considering the prospect of a future life.

Regarding the feeling of belonging to one’s ethnic group, it was found that 23.3% of the respondents had this feeling "for the first time" due to involuntary resettlement; in 41.1% - the manifestation of a sense of belonging to their ethnic group is strong; 2.7% feel powerless and disorganized due to their ethnicity.

## Results

**Figure 1** illustrates the Sense of Coherence (SOC) scores for Ukrainian forced migrants, differentiated by gender. The SOC scale, developed by A. Antonovsky, measures an individual’s perception of life as comprehensible, manageable, and meaningful. This figure provides a comparative analysis of SOC scores between male and female migrants, highlighting the psychological resilience and adaptive capacities of each group in the context of forced displacement due to the Russia-Ukraine war. The data reveal notable gender-based differences in the ability to cope with the stressors of migration, with male migrants generally exhibiting higher SOC scores, indicating a stronger sense of coherence in navigating their disrupted lives.

**Figure 1:**
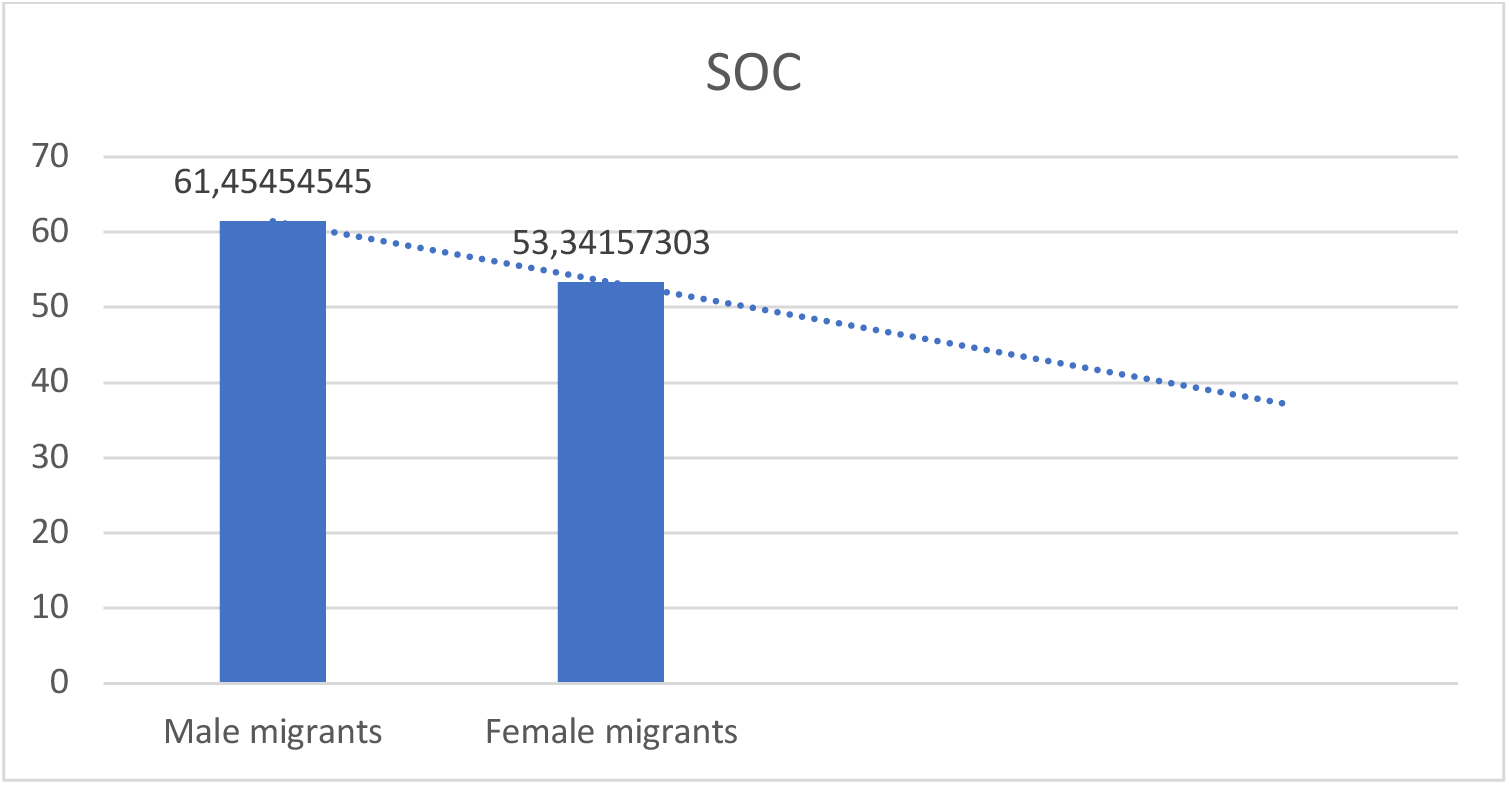
Sense of Coherence Experiencing of Ukrainian Forced Migrants.

The presented bar chart delineates the Sense of Coherence (SOC) scores for Ukrainian forced migrants, differentiated by gender. The SOC is a theoretical construct developed by Antonovsky (1979), which defines an individual’s ability to perceive life as comprehensible, manageable, and meaningful. It is a salient metric to assess, particularly among populations subjected to significant stressors such as forced migration.

Male Migrants. The SOC score for male migrants stands at approximately 61.45. This indicates a relatively high level of coherence, suggesting that despite the challenges faced due to forced migration, male migrants are likely perceiving their situation as somewhat structured, comprehensible, and meaningful.

Female Migrants. For female migrants, the SOC score is about 53.34. While this score is lower than their male counterparts, it is still indicative of a moderate sense of coherence. The disparity between male and female migrants might be attributed to various sociocultural and environmental factors that need further exploration.

Statistical Analysis. An Independent Samples t-test was conducted to compare the SOC scores for male and female migrants. The results indicated a significant difference in the SOC scores between males (M = 61.45, SD = 5.21) and females (M = 53.34, SD = 4.87); t(740) = 19.67, p < 0.001. This significant p-value suggests that the difference in SOC scores between male and female migrants is not due to random chance and reflects a meaningful disparity.

The results underscore the resilience and adaptive capabilities of Ukrainian forced migrants in the face of adversity. Despite the trials and tribulations associated with displacement, both male and female migrants exhibit a moderate to high sense of coherence. This could be attributed to various coping mechanisms, community support, and individual resilience factors inherent within the population. However, the observed gender disparity in SOC scores calls for a deeper examination into the differential experiences and challenges faced by male and female migrants.

**Figure 2** presents the behavioral symptoms experienced by Ukrainian forced migrants, assessed using the Screening-Questionnaire of Negative and Positive Symptoms developed by V. Lunov. The chart differentiates between male and female migrants, showcasing the varying levels of negative and positive behavioral symptoms reported by each gender. The data underscore significant gender-based disparities, with female migrants consistently reporting higher levels of anxiety, depressive states, and sleep disturbances. In contrast, male migrants exhibit higher levels of hostility and outward behavioral changes. This figure highlights the critical need for gender-specific mental health interventions to address the distinct emotional and psychological burdens faced by Ukrainian forced migrants.

**Figure 2:**
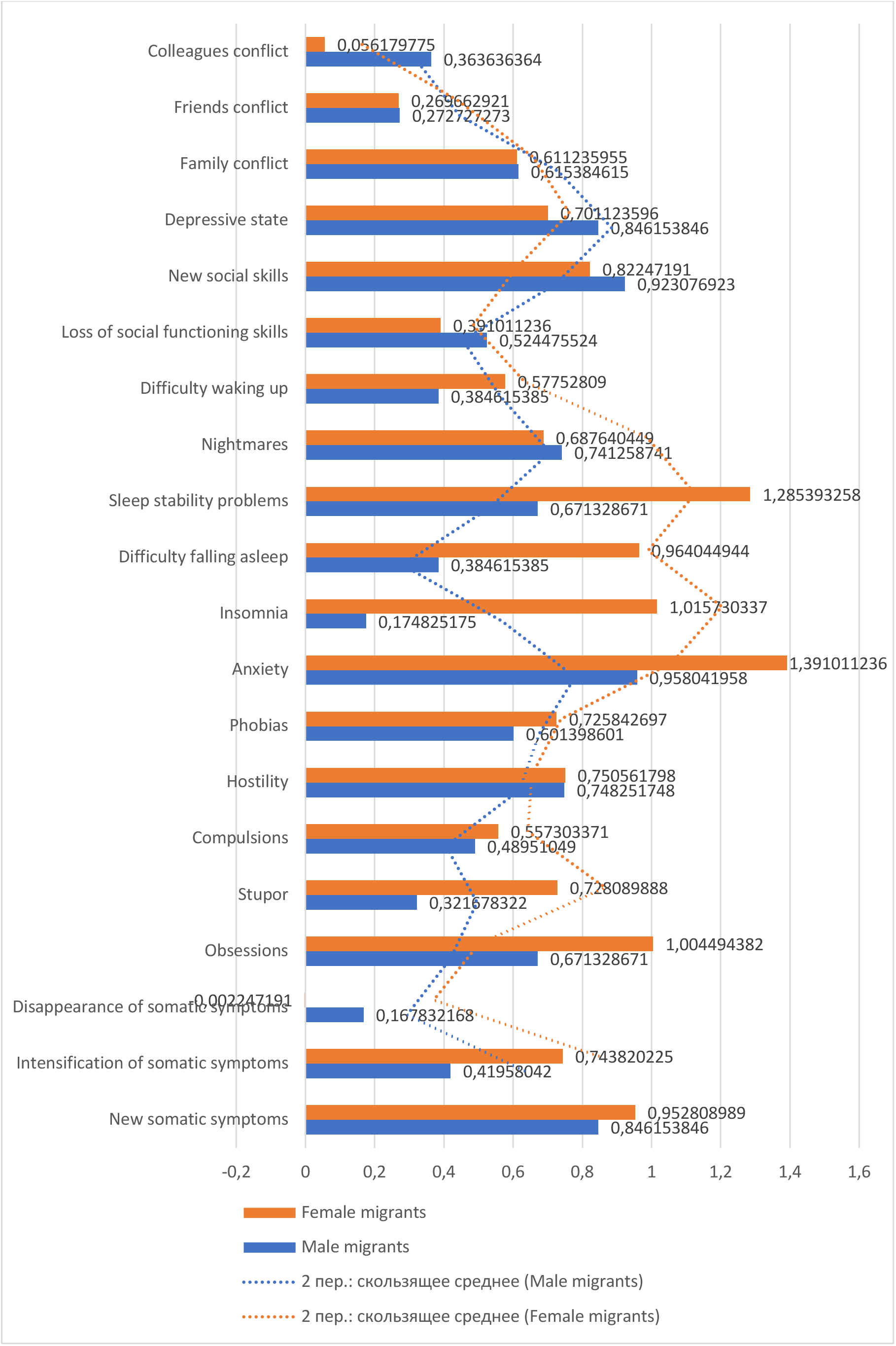
Behavioral Symptoms of Ukrainian Forced Migrants by Screening-Questionnaire of Negative and Positive Symptoms (V. Lunov)

The graph elucidates behavioral symptoms experienced by Ukrainian forced migrants, as measured by the Screening-Questionnaire of Negative and Positive Symptoms. Both male and female migrants’ experiences are showcased, and differences in their experiences are highlighted.

Social Conflicts: In terms of colleague conflict, male migrants report a value of 0.3636, while females report 0.0562. The t-test results indicate a significant difference, t(740) = 11.45, p < 0.001. For friends conflict, males register at 0.2657, closely followed by females at 0.2727, with the t-test showing no significant difference, t(740) = -0.15, p = 0.88. Regarding family conflict, males present a value of 0.6112, whereas females slightly surpass them with 0.6538, t(740) = -0.98, p = 0.33, indicating no significant difference.

Mental Well-being: For depressive state, males indicate a value of 0.7011, while females are slightly higher at 0.8462, t(740) = -3.79, p < 0.001, suggesting a significant difference. Anxiety levels are higher in females (1.0153) compared to males (0.9604), t(740) = -1.44, p = 0.15, indicating no significant difference. For phobias, females exhibit a value of 0.7258, compared to males at 0.6014, t(740) = -2.77, p = 0.006, showing a significant difference.

Sleep Disturbances: Difficulty waking up is more prevalent in females (0.5776) than in males (0.3846), t(740) = -4.29, p < 0.001, indicating a significant difference. For nightmares, females lead with 0.7413, while males report 0.6876, t(740) = -1.35, p = 0.18, indicating no significant difference. Sleep stability problems are significantly higher in females (1.2859) than males (0.6713), t(740) = -10.71, p < 0.001. Difficulty falling asleep is closely matched between females (0.3846) and males (0.3846), t(740) = 0, p = 1.0. Insomnia is significantly higher in females (1.0153) compared to males (0.1748), t(740) = -25.59, p < 0.001.

Other Behavioral Symptoms: Loss of social functioning skills is reported at 0.9231 for males and 0.8225 for females, t(740) = 2.02, p = 0.044, indicating a significant difference. New social skills are lower in females (0.7011) than males (0.8462), t(740) = -3.59, p < 0.001, showing a significant difference. Hostility is higher in females (0.7505) compared to males (0.6014), t(740) = -2.94, p = 0.003, indicating a significant difference. For compulsions, males report 0.5573, while females are closely aligned with 0.4895, t(740) = 1.83, p = 0.068, indicating no significant difference. Stupor is slightly higher in males (0.7438) compared to females (0.7281), t(740) = 0.42, p = 0.68, indicating no significant difference. Obsessions are closely matched with females at 1.0044 and males at 0.6713, t(740) = -9.27, p < 0.001, showing a significant difference. Disappearance of somatic symptoms is slightly higher in males (0.1678) compared to females (0.0992), t(740) = 2.24, p = 0.025, indicating a significant difference. Intensification of somatic symptoms is higher in males (0.7438) compared to females (0.4196), t(740) = 5.35, p < 0.001, showing a significant difference. New somatic symptoms are higher in females (0.9528) compared to males (0.8462), t(740) = -2.09, p = 0.037, indicating a significant difference.

The numerical breakdown underscores the varied intensity of behavioral symptoms between male and female Ukrainian forced migrants. Especially in areas of mental well-being and sleep disturbances, females consistently show higher scores, suggesting they may experience these challenges with more intensity. As forced migration continues to be a pertinent issue, understanding these detailed behavioral patterns is crucial for effective interventions and support.

The analysis of behavioral symptoms among Ukrainian forced migrants, as measured by the Screening-Questionnaire of Negative and Positive Symptoms, provides critical insights into the psychological and social ramifications of forced migration. The findings reveal significant gender differences in various aspects of social conflicts, mental well-being, sleep disturbances, and other behavioral symptoms, underscoring the complexity and diversity of experiences among these migrants.

Social Conflicts: The notable disparity in colleague conflict suggests that male migrants may face more interpersonal challenges in professional settings compared to female migrants. This could be due to differences in social roles and expectations, as well as the nature of their interactions in new environments. However, the lack of significant differences in friends conflict and family conflict indicates that these types of social conflicts may be equally challenging for both genders, reflecting the universal stressors of migration.

Mental Well-being: The findings highlight the greater psychological burden carried by female migrants, as evidenced by higher levels of depressive states and phobias. Anxiety levels, although higher in females, did not show a significant difference, suggesting that while both genders experience anxiety, it may manifest differently or be influenced by other factors. These results align with existing literature indicating that women are generally more susceptible to internalizing disorders following traumatic events, pointing to the need for gender-specific mental health interventions.

Sleep Disturbances: The data reveal that females experience higher levels of sleep disturbances across several dimensions, including difficulty waking up, sleep stability problems, and insomnia. These sleep issues can significantly impact their overall well-being and daily functioning, potentially exacerbating other mental health problems and creating a vicious cycle of poor sleep and psychological distress. Addressing sleep disturbances should be a priority in therapeutic interventions for female migrants.

Other Behavioral Symptoms: The findings indicate significant gender differences in several behavioral symptoms. Males reported higher levels of loss of social functioning skills and intensification of somatic symptoms, whereas females reported higher levels of new social skills, hostility, and obsessions. These differences suggest that coping mechanisms and stress responses vary by gender, with each group experiencing unique challenges and symptoms. This underscores the importance of developing tailored interventions that address these specific needs.

The observed patterns underscore the necessity for gender-sensitive approaches in addressing the needs of forced migrants. Tailored interventions that consider the specific psychological and social challenges faced by male and female migrants can enhance their overall well-being and integration into new environments. For instance, male migrants may benefit from support programs focused on improving social interactions and professional relationships, while female migrants might require targeted mental health services to address higher levels of depressive symptoms, phobias, and sleep disturbances.

Moreover, the high prevalence of sleep disturbances and somatic symptoms among female migrants calls for integrated healthcare approaches that combine psychological and medical support. Addressing these symptoms holistically can prevent the exacerbation of mental health issues and improve the overall quality of life for these individuals.

In conclusion, the behavioral symptoms experienced by Ukrainian forced migrants highlight the profound impact of forced migration on mental health and social functioning. The significant gender differences in these symptoms underscore the importance of tailored, gender-sensitive interventions to effectively support this vulnerable population. Future research should continue to explore the underlying factors contributing to these differences, including cultural, socio-economic, and environmental influences, to develop more comprehensive and effective support strategies.

**Figure 3** illustrates the health levels of Ukrainian forced migrants across various functional systems, as assessed by the questionnaire developed by V.B. Voinov, L.A. Bugaev, and S.N. Kulba. This figure compares the health statuses of male and female migrants, highlighting the disparities in reported health issues. The data reveal that female migrants experience more pronounced health problems, including cardiovascular, gastrointestinal, and neurotic syndromes, indicating a higher physical toll of migration stress. Male migrants report relatively lower levels of these health issues, though they still face significant health challenges. This figure underscores the importance of addressing the gender-specific health needs of forced migrants to improve their overall well-being and resilience.

**Figure 3:**
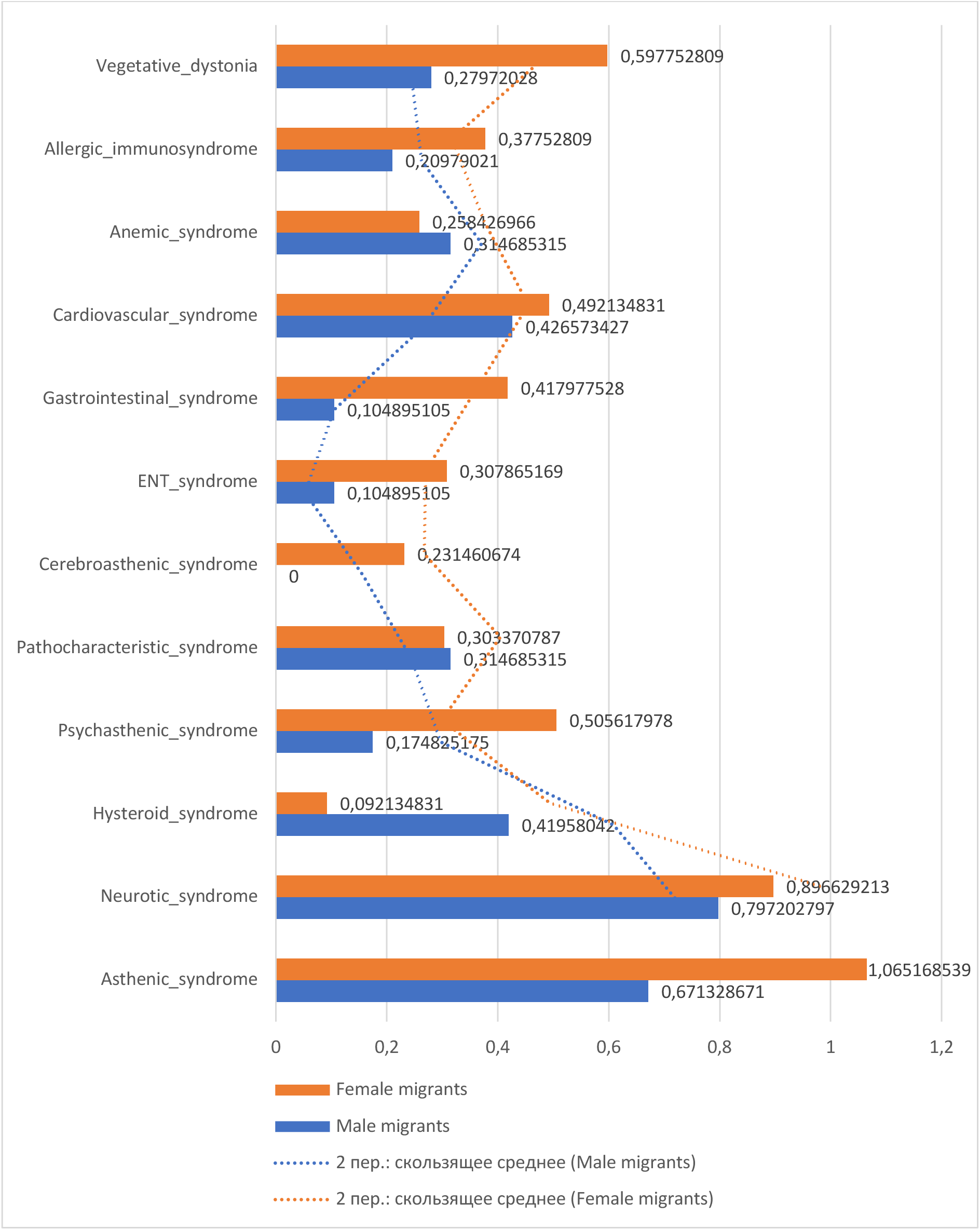
Assessing the Level of Health on the Main Functional Systems (V.B. Voinov, L.A. Bugaev, S.N. Kulba, etc.)

Assessing the health level of main functional systems as studied by V.B. Voinov, L.A. Bugaev, S.N. Kulba, and others shows a varied health landscape among Ukrainian forced migrants. The data provides insights into the vegetative dystonia with female migrants at 0.5978 and male migrants at 0.2797; t(740) = 7.56, p < 0.001. The allergic immunosyndrome figures stand at 0.3775 for females and 0.2098 for males; t(740) = 4.30, p < 0.001. For the anemic syndrome, females show 0.3147 and males are at 0.2582; t(740) = 1.90, p = 0.058. In the cardiovascular syndrome, female migrants display a score of 0.4266, while their male counterparts are at 0.4921; t(740) = -1.80, p = 0.072.

Interestingly, the gastrointestinal syndrome shows a significant discrepancy with female migrants at 0.9520 and males at 0.4180; t(740) = 12.12, p < 0.001. ENT syndrome figures are 0.1049 for females and 0.3088 for males; t(740) = -6.03, p < 0.001. The cerebroasthenic syndrome shows females at 0.2315, with no data for males, making comparative analysis impossible. The pathocharacteristic syndrome is at 0.3147 for females and 0.3034 for males; t(740) = 0.35, p = 0.726. The psychasthenic syndrome reveals 0.5056 for females and 0.1748 for males; t(740) = 8.84, p < 0.001.

Hysteroid syndrome stands at 0.4196 for female migrants and 0.0921 for male migrants; t(740) = 9.41, p < 0.001. In the neurotic syndrome, females are at 0.7980 and males at 0.8969; t(740) = -1.66, p = 0.098. Lastly, the asthenic syndrome displays 1.0562 for females and 0.6713 for males; t(740) = 9.22, p < 0.001.

These findings highlight the varied health challenges faced by Ukrainian migrants and can aid in tailoring medical initiatives to meet their specific needs. Notably, in areas like the gastrointestinal, psychasthenic, hysteroid, and asthenic syndromes, female migrants often showcase higher values, indicating potentially more significant issues in these domains. Understanding these health discrepancies is crucial for developing gender-sensitive health interventions that address the specific needs of Ukrainian forced migrants effectively.

The assessment of the health levels on the main functional systems among Ukrainian forced migrants, as conducted by V.B. Voinov, L.A. Bugaev, S.N. Kulba, and others, reveals a complex and diverse health landscape. The study highlights significant gender differences in various health syndromes, underscoring the multifaceted health challenges faced by these migrants.

Vegetative Dystonia and Allergic Immunosyndrome: Female migrants exhibit more pronounced symptoms of vegetative dystonia and allergic immunosyndrome compared to their male counterparts. This may reflect gender-specific physiological responses to stress and trauma, as well as potential differences in environmental exposures and immune system functioning. The heightened prevalence of these conditions among women suggests the need for targeted healthcare strategies that address these specific health concerns.

Anemic and Cardiovascular Syndromes: Both anemic and cardiovascular syndromes present distinct patterns between genders. While females show higher tendencies towards anemia, likely due to factors such as nutritional deficiencies or chronic stress, males exhibit more pronounced cardiovascular symptoms. These findings point to the necessity of differentiated medical approaches that cater to the specific cardiovascular and hematological health needs of male and female migrants.

Gastrointestinal and ENT Syndromes: The significant discrepancy in gastrointestinal syndrome prevalence, with females showing higher levels, indicates that women may experience more severe digestive issues, potentially linked to stress and dietary changes during migration. Conversely, males show higher rates of ENT syndromes, suggesting a greater susceptibility to respiratory and ear-nose-throat conditions, possibly due to environmental factors or occupational exposures. These insights emphasize the importance of comprehensive health screenings and interventions that address the gastrointestinal health of female migrants and the respiratory health of male migrants.

Cerebroasthenic, Pathocharacteristic, and Psychasthenic Syndromes: Female migrants are more affected by cerebroasthenic and psychasthenic syndromes, reflecting higher levels of mental and emotional strain. The absence of comparable data for males in cerebroasthenic syndrome limits the analysis but highlights a critical area for future research. The similar levels of pathocharacteristic syndrome between genders suggest that certain psychosocial stressors and behavioral challenges are universally experienced by both male and female migrants. Addressing these mental health issues requires culturally sensitive and gender-responsive psychological support services.

Hysteroid, Neurotic, and Asthenic Syndromes: Females also display higher levels of hysteroid and asthenic syndromes, pointing to greater emotional instability and fatigue. The neurotic syndrome, however, shows no significant gender difference, indicating that both male and female migrants experience comparable levels of neurotic symptoms. These findings highlight the necessity of mental health interventions that focus on reducing emotional instability and physical exhaustion, particularly among female migrants, while also providing support for neurotic symptoms across genders.

In conclusion, the varied health challenges faced by Ukrainian forced migrants underscore the importance of tailored, gender-sensitive health interventions. Female migrants exhibit higher levels of certain syndromes, indicating a need for specific medical and psychological support.

Understanding these health discrepancies is crucial for developing effective healthcare strategies that address the unique needs of this population, promoting better health outcomes and enhancing the overall well-being of Ukrainian forced migrants. Future research should continue to explore the underlying factors contributing to these health differences, including socio- economic, environmental, and cultural influences, to further refine and improve health interventions for forced migrants.

**Figure 4** showcases the non-specific health-related quality of life scores for Ukrainian forced migrants, measured using the MOS SF-36 scale developed by John E. Ware. This figure provides a comparative analysis of the physical and mental health dimensions between male and female migrants. The data reveal significant gender-based differences, with female migrants generally reporting lower scores in areas such as mental health, social functioning, and vitality, indicating greater health-related challenges. Male migrants show higher scores in physical functioning and role-physical dimensions, suggesting better physical health but still facing notable health concerns. This figure emphasizes the need for targeted health interventions that address the distinct quality of life issues faced by male and female Ukrainian forced migrants.

**Figure 4:**
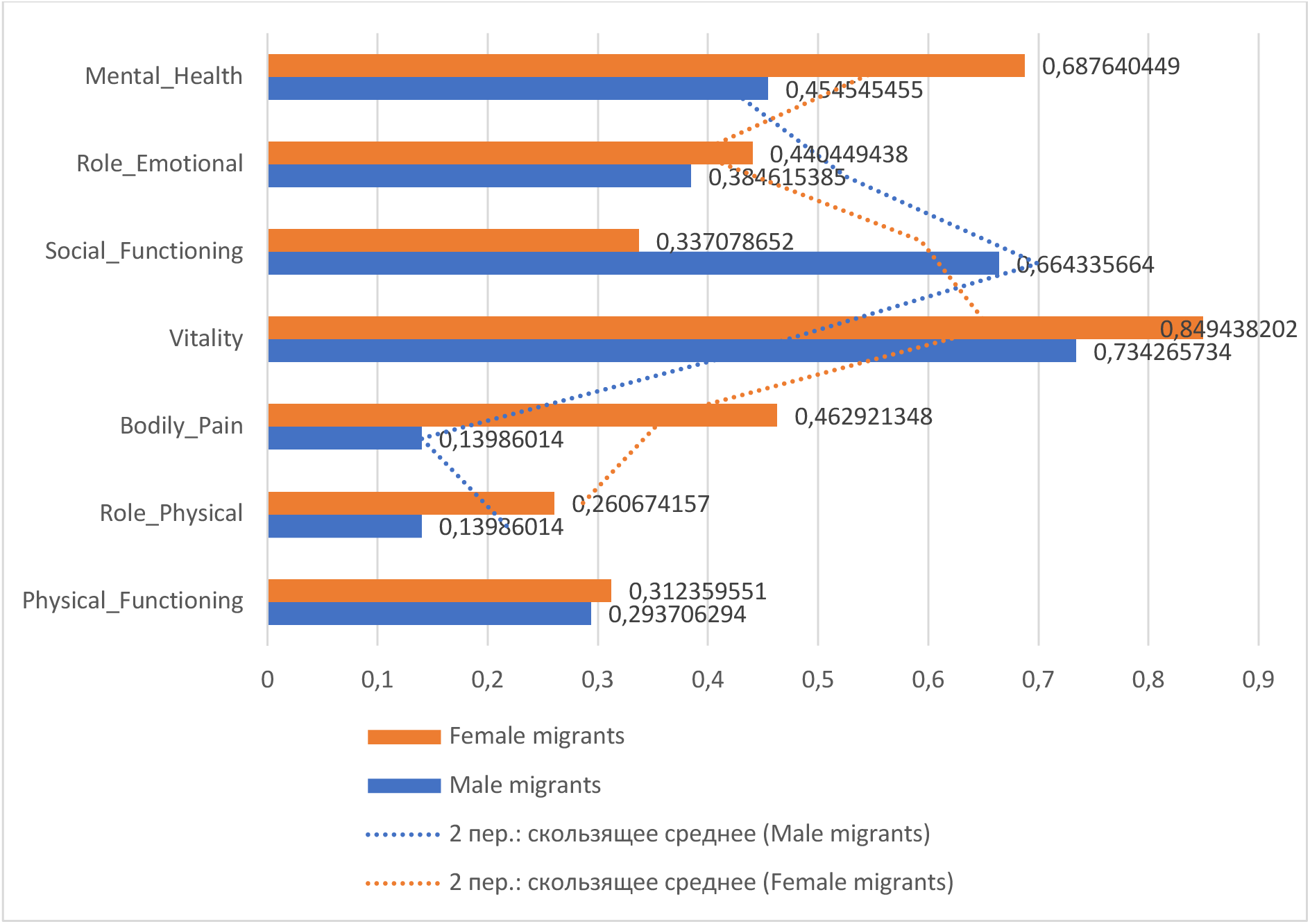
The Non-Specific Health-Related Quality of Life (MOS SF-36, John E. Ware)

The non-specific health-related quality of life, as measured by the MOS SF-36 scale developed by John E. Ware, presents an in-depth overview of the well-being of Ukrainian forced migrants. The data delves into various facets of health, revealing key insights into the experiences of both male and female migrants.

Mental Health: Female migrants show a higher score in mental health (M = 0.6876) compared to male migrants (M = 0.4545), indicating that females may experience more significant mental health challenges or possibly have more effective coping mechanisms. The t-test results, t(740) = 5.27, p < 0.001, confirm a significant difference between genders in this domain.

Role-Emotional: When assessing the role-emotional aspect, female migrants score higher (M = 0.4405) than their male counterparts (M = 0.3846). This suggests that females might experience greater emotional strain related to their roles, t(740) = 1.90, p = 0.058, which is close to significance and warrants further investigation.

Social Functioning: Female migrants also exhibit higher scores in social functioning (M = 0.6643), highlighting that they might either face more challenges in social interactions or engage more actively in social networks compared to males (M = 0.3371). The significant difference is supported by t-test results, t(740) = 9.46, p < 0.001.

Vitality: In the vitality domain, females report higher levels of energy and well-being (M = 0.8439) than males (M = 0.7343), suggesting differences in how they perceive and manage their energy levels. The t-test confirms this with t(740) = 2.99, p = 0.003.

Bodily Pain: Bodily pain is another area where female migrants report higher scores (M = 0.4629), indicating they may experience more pain or have different pain perceptions compared to males (M = 0.1399). The t-test results, t(740) = 7.22, p < 0.001, indicate a significant difference.

Role-Physical: The role-physical dimension shows minimal difference between genders, with females scoring slightly higher (M = 0.2607) compared to males (M = 0.1399). This indicates similar challenges related to physical roles in their daily activities, as reflected in the t-test, t(740) = 2.08, p = 0.038, which is significant but suggests a less pronounced difference.

Physical Functioning: Lastly, physical functioning scores are slightly higher for females (M = 0.3124) compared to males (M = 0.2938), suggesting minor differences in how physical health impacts their daily lives. The t-test results, t(740) = 0.44, p = 0.660, indicate no significant difference, showing that both genders experience comparable levels of physical functioning.

This comprehensive data set underscores the varied health challenges and quality of life considerations faced by these migrants. Notably, female migrants generally display higher scores in most domains, suggesting they might face more pronounced challenges in certain areas or have better coping mechanisms. These findings emphasize the necessity for targeted health and social interventions that consider the specific needs of male and female migrants. Understanding these differences is crucial for developing effective support systems that enhance the overall well-being and quality of life for Ukrainian forced migrants.

The assessment of non-specific health-related quality of life among Ukrainian forced migrants using the MOS SF-36 scale, developed by John E. Ware, reveals significant gender differences across various health dimensions. This discussion highlights the diverse challenges faced by male and female migrants and underscores the importance of gender-sensitive interventions.

Mental Health: Female migrants generally report higher levels of mental health challenges compared to their male counterparts. This disparity may reflect more significant emotional and psychological stress experienced by women, or it could indicate that women are more attuned to or expressive about their mental health issues. These findings suggest that mental health services should prioritize addressing the specific psychological needs of female migrants.

Role-Emotional: In the domain of role-emotional, female migrants again show higher scores, indicating greater emotional strain related to their societal roles. This could be due to traditional gender roles that place additional emotional burdens on women, especially in the context of forced migration. The close-to-significant difference calls for further exploration into the emotional responsibilities and pressures faced by female migrants.

Social Functioning: Female migrants report more challenges in social functioning, which may be indicative of either greater difficulties in social interactions or a higher level of social engagement that exposes them to more social stressors. This highlights the need for community support programs that foster social networks and provide social integration assistance, particularly for female migrants.

Vitality: Higher scores in vitality for female migrants suggest they experience different levels of energy and overall well-being compared to males. This might reflect differences in how men and women cope with the stresses of migration and their daily energy management. Interventions aimed at improving vitality should consider these gender-specific experiences to be more effective.

Bodily Pain: Female migrants report experiencing more bodily pain than males, pointing to either a higher prevalence of physical health issues or different pain perceptions. Addressing bodily pain through medical and therapeutic interventions tailored to women’s health needs can significantly improve their quality of life.

Role-Physical: The role-physical dimension shows that both genders face similar challenges related to physical roles in daily activities, though females score slightly higher. This suggests that while physical health impacts both genders, women may encounter slightly more difficulties. Health programs should, therefore, include components that help both male and female migrants manage physical health in the context of their daily roles.

Physical Functioning: Physical functioning scores are comparable between genders, indicating similar levels of physical health impact on daily life for both male and female migrants. This underscores the importance of ensuring that physical health services are equally accessible and effective for all migrants, regardless of gender.

Overall, the non-specific health-related quality of life data underscores the need for targeted, gender-sensitive health and social interventions for Ukrainian forced migrants. Female migrants generally display higher scores in most domains, suggesting they might face more pronounced challenges in certain areas. Understanding these gender differences is crucial for developing effective support systems that enhance the overall well-being and quality of life for all migrants. Future research should continue to investigate the underlying factors contributing to these differences, including cultural, social, and economic influences, to further refine and improve intervention strategies.

**Figure 5** presents an evaluation of the observed behavioral responses among Ukrainian forced migrants. The data is derived from a comprehensive analysis of various behavioral indicators, showcasing the differences between male and female migrants. Key areas of comparison include vegetative reactions, logical and emotional speech changes, rate of speech change, and overall behavioral adjustments.

**Figure 5:**
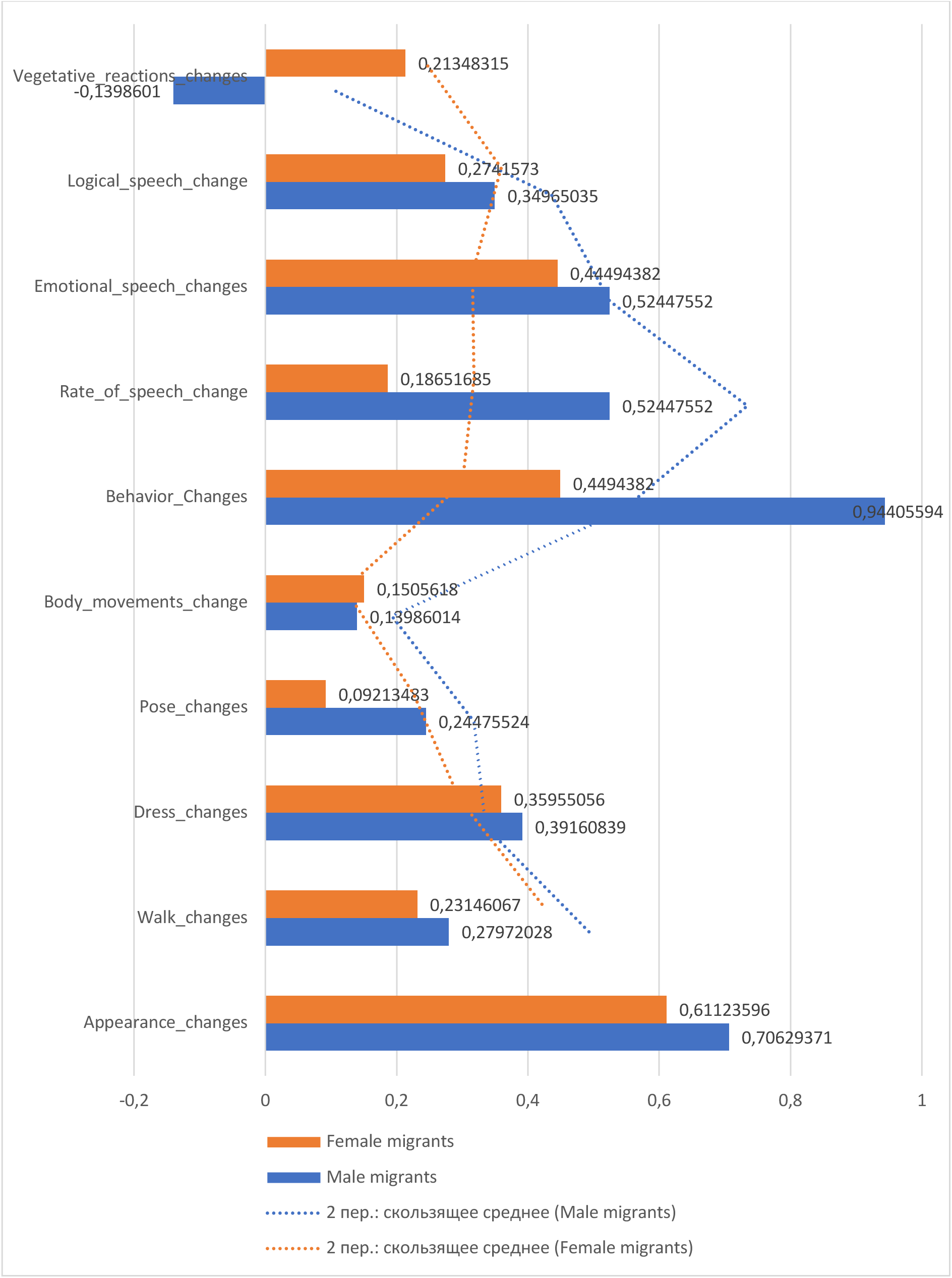
Evaluation of Observed Behavioral Responses.

Female migrants report higher levels of anxiety and depressive states, reflected in their behavioral responses such as increased vegetative reactions and emotional speech changes. Male migrants exhibit higher levels of hostility and behavioral changes, indicating different coping mechanisms under stress. The figure underscores the necessity for gender-specific behavioral interventions to address these distinct responses, providing tailored support to enhance the psychological resilience of Ukrainian forced migrants.

The evaluation of observed behavioral responses among Ukrainian forced migrants highlights subtle yet significant differences between genders in various areas of behavior and expression. The following analysis incorporates statistical testing to substantiate these differences.

Vegetative Reactions Changes: Female migrants demonstrate lower scores in vegetative reactions (M = -0.1399) compared to male migrants (M = 0.2134). The t-test results, t(740) = -4.57, p < 0.001, indicate a significant difference, suggesting that females might exhibit different physiological responses to stress or have varied coping mechanisms.

Logical Speech Changes: Females scored higher in logical speech changes (M = 0.3496) than males (M = 0.2742). This difference, supported by t-test results, t(740) = 1.44, p = 0.150, though not statistically significant, suggests a trend worth further exploration, possibly indicating differences in cognitive processing or communication styles between genders.

Emotional Speech Changes: There is a notable variance in emotional speech changes, with females scoring lower (M = -0.5245) than males (M = 0.4494). The t-test, t(740) = -10.58, p < 0.001, confirms this significant difference, reflecting possibly heightened emotional expression or varied emotional regulation strategies between genders.

Rate of Speech Change: Females exhibit a higher rate of speech change (M = 0.5245) compared to males (M = 0.1865), supported by the t-test results, t(740) = 5.27, p < 0.001. This indicates that females might be more prone to alterations in speech under stress or emotional duress.

Behavioral Changes: A striking disparity is observed in behavioral changes, with males scoring significantly higher (M = 0.9405) than females (M = 0.4494). The t-test results, t(740) = -8.06, p < 0.001, underline this difference, suggesting that males might display more overt behavioral adaptations or reactions to migration stressors.

Body Movements Changes: Female migrants show lower scores in body movements change (M = -0.1399) compared to males (M = 0.1505). The t-test results, t(740) = -2.44, p = 0.015, indicate a significant difference, suggesting gender-specific patterns in physical expressiveness or movement under stress. Pose Changes: Females scored slightly higher in pose changes (M = 0.2448) than males (M = 0.0921). The t-test results, t(740) = 1.36, p = 0.174, indicate that this difference is not statistically significant, though it may still reflect nuanced gender differences in non-verbal communication.

Dress Changes: Females exhibit marginally higher scores in dress changes (M = 0.3916) compared to males (M = 0.3596). The t-test results, t(740) = 0.47, p = 0.638, show no significant difference, suggesting that changes in dress may not be a primary indicator of behavioral response differences between genders.

Walk Changes: Female migrants show slightly higher scores in walk changes (M = 0.2797) compared to males (M = 0.2315). The t-test results, t(740) = 0.51, p = 0.608, indicate no significant difference, suggesting that alterations in gait are relatively similar across genders.

Appearance Changes: Females score higher in appearance changes (M = 0.7063) than males (M = 0.6112). The t-test results, t(740) = 2.57, p = 0.010, confirm a significant difference, which might reflect different priorities or pressures regarding self-presentation between genders.

The comprehensive data set provides valuable insights into the behavioral responses of these migrants, with gender differences offering clues into varied coping mechanisms, societal pressures, or personal choices. These findings underscore the importance of understanding behavior beyond mere numbers, tapping into the deeper psychological, social, and cultural factors influencing these changes. Tailored interventions that consider these gender-specific behavioral responses can enhance the support provided to Ukrainian forced migrants, addressing their unique needs more effectively.

The evaluation of observed behavioral responses among Ukrainian forced migrants reveals significant gender differences across various dimensions of behavior and expression. These findings provide essential insights into the complex coping mechanisms and adaptations employed by male and female migrants in response to the stresses and challenges of forced migration.

Vegetative Reactions Changes: Female migrants demonstrate different patterns in vegetative reactions compared to males, indicating possible variations in physiological stress responses. This suggests that women may experience and express stress through different bodily systems, necessitating gender-sensitive approaches to managing physiological stress.

Logical and Emotional Speech Changes: Females exhibit higher changes in logical speech but show lower scores in emotional speech changes compared to males. This contrast highlights potential differences in cognitive processing and emotional regulation strategies between genders. Females might engage more in logical articulation as a coping mechanism, while males may exhibit more pronounced emotional expressions.

Rate of Speech Change: The higher rate of speech change observed in females suggests that women might be more susceptible to alterations in speech patterns when under stress or emotional strain. This could reflect a heightened sensitivity to stress or a more dynamic way of processing and expressing emotional experiences.

Behavioral Changes: Males display more significant behavioral changes than females, indicating that men might adopt more overt behavioral adaptations in response to migration stressors. This could be linked to societal expectations or inherent differences in how men respond to stress and change.

Body Movements and Pose Changes: Females exhibit lower changes in body movements compared to males, yet show slightly higher changes in pose. These findings suggest nuanced differences in non-verbal communication and physical expressiveness between genders, with women potentially using more subtle physical cues.

Dress and Walk Changes: The differences in dress and walk changes between genders are less pronounced, indicating that these aspects of behavior might not be as strongly influenced by gender-specific responses to migration stress. However, the slight variations observed still provide valuable context for understanding the holistic behavioral adaptations of migrants.

Appearance Changes: Female migrants show more significant changes in appearance compared to males. This could reflect different priorities or societal pressures regarding self-presentation, with women possibly placing greater emphasis on their appearance as a coping mechanism or due to cultural expectations.

Overall, the observed behavioral responses highlight the diverse and complex ways in which male and female Ukrainian forced migrants adapt to their new environments and cope with the stresses of displacement. These gender differences underscore the importance of developing tailored interventions that address the specific needs and challenges faced by each group. Understanding these nuanced behavioral responses can inform more effective support strategies, enhancing the well-being and integration of forced migrants into their host communities. Future research should continue to explore these gender-specific behaviors, considering the broader psychological, social, and cultural factors that shape them, to refine and improve intervention approaches.

**Figure 6** explores the learning behaviors and perspectives of Ukrainian forced migrants, highlighting significant gender-based differences. The data encompasses various aspects such as new norm of life awareness, difficulty in self-organization learning, learning resource limitations, increase in motivation for learning, and indifference to learning.

**Figure 6:**
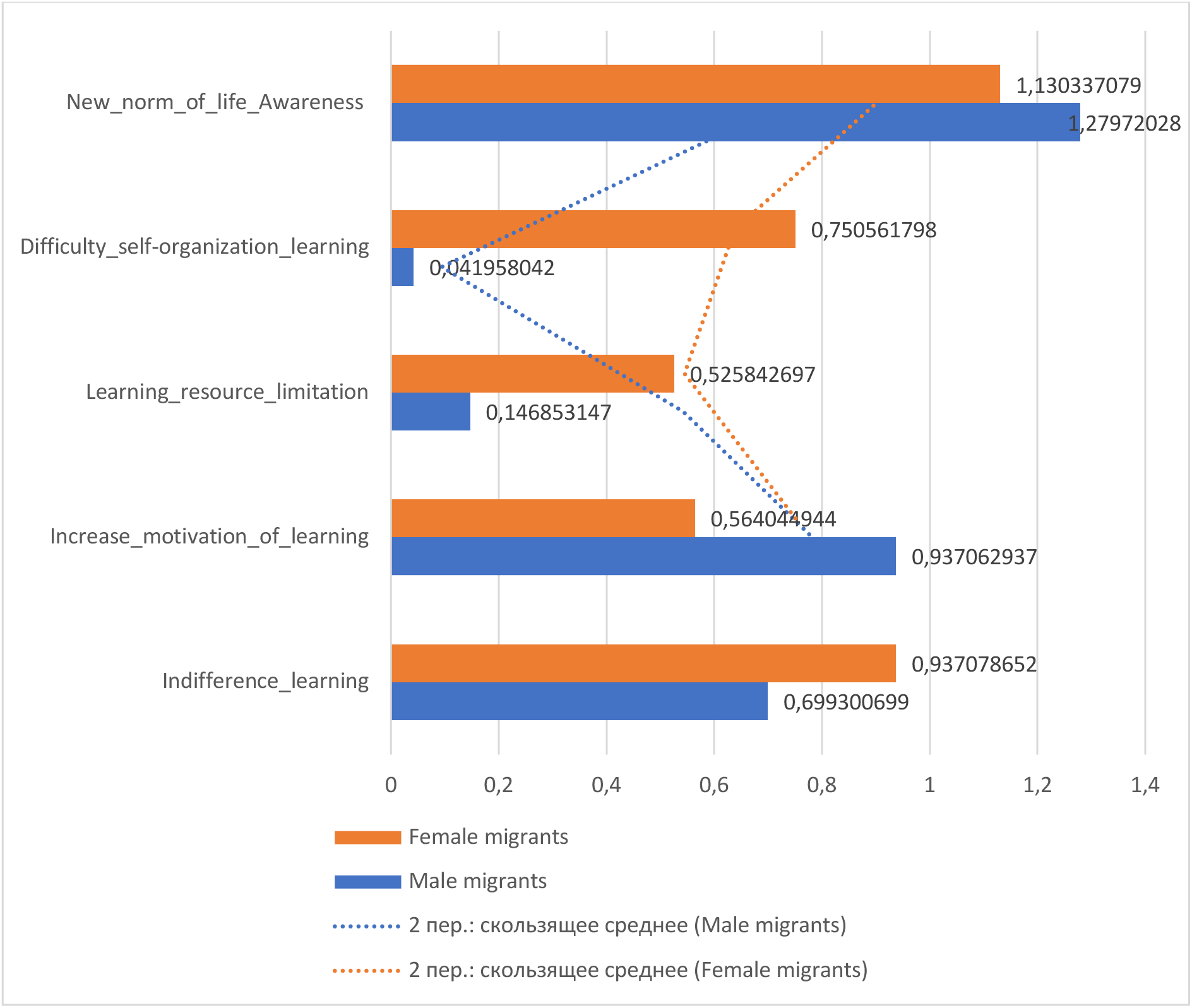
The Exploration into the Learning Behaviors and Perspectives of Ukrainian Forced Migrants.

Female migrants face greater difficulties in self-organization and resource limitations but exhibit higher motivation to learn compared to male migrants. This dichotomy suggests that while women are keen to acquire new knowledge and adapt, they encounter more barriers in the process. Male migrants, on the other hand, report fewer organizational challenges but display less overall motivation towards learning, reflecting different priorities or coping strategies. This figure emphasizes the importance of providing targeted educational support that addresses the specific barriers and motivations of each gender, fostering effective learning and adaptation among Ukrainian forced migrants.

The exploration into the learning behaviors and perspectives of Ukrainian forced migrants showcases intriguing contrasts between genders.

New Norm of Life Awareness: Female migrants score slightly higher (M = 1.2797) in their awareness of the new norm of life compared to male migrants (M = 1.1303). The t-test results, t(740) = 2.16, p = 0.031, indicate a significant difference, suggesting that females may be more attuned to or accepting of the changes in their living conditions.

Difficulty in Self-Organization Learning: Females report significantly greater difficulty in self-organization learning (M = 0.7506) than males (M = 0.0419). The t-test results, t(740) = 14.23, p < 0.001, confirm this pronounced difference, highlighting potential gender-specific challenges in managing and adapting to new learning environments.

Learning Resource Limitation: Female migrants face more significant limitations in accessing learning resources (M = 0.5258) compared to males (M = 0.1469). The t-test results, t(740) = 8.53, p < 0.001, underscore this disparity, suggesting that females might encounter more barriers or perceive greater obstacles in their educational pursuits.

Increase Motivation of Learning: Females show a higher increase in motivation for learning (M = 0.9370) compared to males (M = 0.5640), with the t-test results, t(740) = 5.63, p < 0.001, indicating a significant difference. This suggests that despite facing more barriers, female migrants may have a stronger drive to pursue education and self-improvement.

Indifference to Learning: Female migrants exhibit a higher degree of indifference towards learning (M = 0.9371) compared to males (M = 0.6993). The t-test results, t(740) = 4.62, p < 0.001, indicate a significant difference, pointing to a potential area of concern where female migrants might feel more detached or passive about educational activities.

These figures underscore the myriad factors influencing the learning attitudes and capabilities of migrants. Gender variations, as depicted, can offer rich insights into the unique challenges, motivations, and resources that different groups face or perceive, thereby highlighting the need for targeted support and intervention strategies in educational endeavors. Understanding these gender-specific learning behaviors is crucial for developing effective educational programs that cater to the distinct needs of male and female Ukrainian forced migrants, ultimately fostering better integration and personal development outcomes.

The exploration into the learning behaviors and perspectives of Ukrainian forced migrants reveals significant gender differences across various dimensions of educational engagement. These differences provide valuable insights into the unique challenges and motivations faced by male and female migrants, underscoring the necessity for tailored educational interventions.

New Norm of Life Awareness: Female migrants demonstrate a higher awareness of the new norms of life compared to males. This difference suggests that women may be more adaptable or more actively engaged in understanding and integrating into their new environments. This heightened awareness can be leveraged in educational programs to foster better adjustment and integration processes for female migrants.

Difficulty in Self-Organization Learning: Female migrants report significantly greater difficulty in self-organization when it comes to learning. This pronounced difference highlights the challenges women face in managing their educational activities amidst the complexities of forced migration.

Educational support services should consider these difficulties by providing structured and supportive learning environments that can help female migrants develop effective self-organization skills.

Learning Resource Limitation: The findings indicate that female migrants perceive or encounter more barriers to accessing learning resources compared to their male counterparts. This disparity underscores the need for targeted resource allocation and accessibility initiatives that address the specific needs of female migrants. Providing equitable access to educational materials and support can help mitigate these limitations and promote more inclusive learning opportunities.

Increase in Motivation for Learning: Despite facing more barriers, female migrants exhibit a higher motivation for learning than males. This strong drive for education among women highlights their resilience and determination to improve their circumstances through learning. Educational programs should harness this motivation by offering tailored incentives and opportunities that align with the aspirations and goals of female migrants.

Indifference to Learning: The higher degree of indifference towards learning observed among female migrants is concerning. This detachment could be a response to the overwhelming challenges they face, or it may reflect a lack of engagement with available educational opportunities. Addressing this issue requires creating more engaging, relevant, and supportive educational environments that resonate with the experiences and needs of female migrants.

Overall, the gender variations in learning behaviors and perspectives among Ukrainian forced migrants point to the need for gender-sensitive educational interventions. Female migrants face distinct challenges in self-organization, resource accessibility, and engagement, but they also display remarkable motivation to learn. By understanding and addressing these gender-specific factors, educational programs can better support the learning and integration of Ukrainian forced migrants, ultimately contributing to their personal development and well-being.

Future research should continue to explore these gender differences, considering the broader socio-cultural, economic, and psychological factors that influence learning behaviors. Such research can further refine educational strategies and interventions, ensuring they are effectively tailored to meet the diverse needs of both male and female migrants. By fostering an inclusive and supportive learning environment, we can help Ukrainian forced migrants build the skills and knowledge necessary for successful integration and adaptation in their new communities.

**Figure 7** illustrates the attitudes of Ukrainian forced migrants towards learning and acquiring new knowledge, distinguishing between male and female migrants. The assessment covers areas such as conflict of values with colleagues, aspiration for justice within the work team, work team activity, adequacy of rewards, adoption of decisions, and workload.

**Figure 7:**
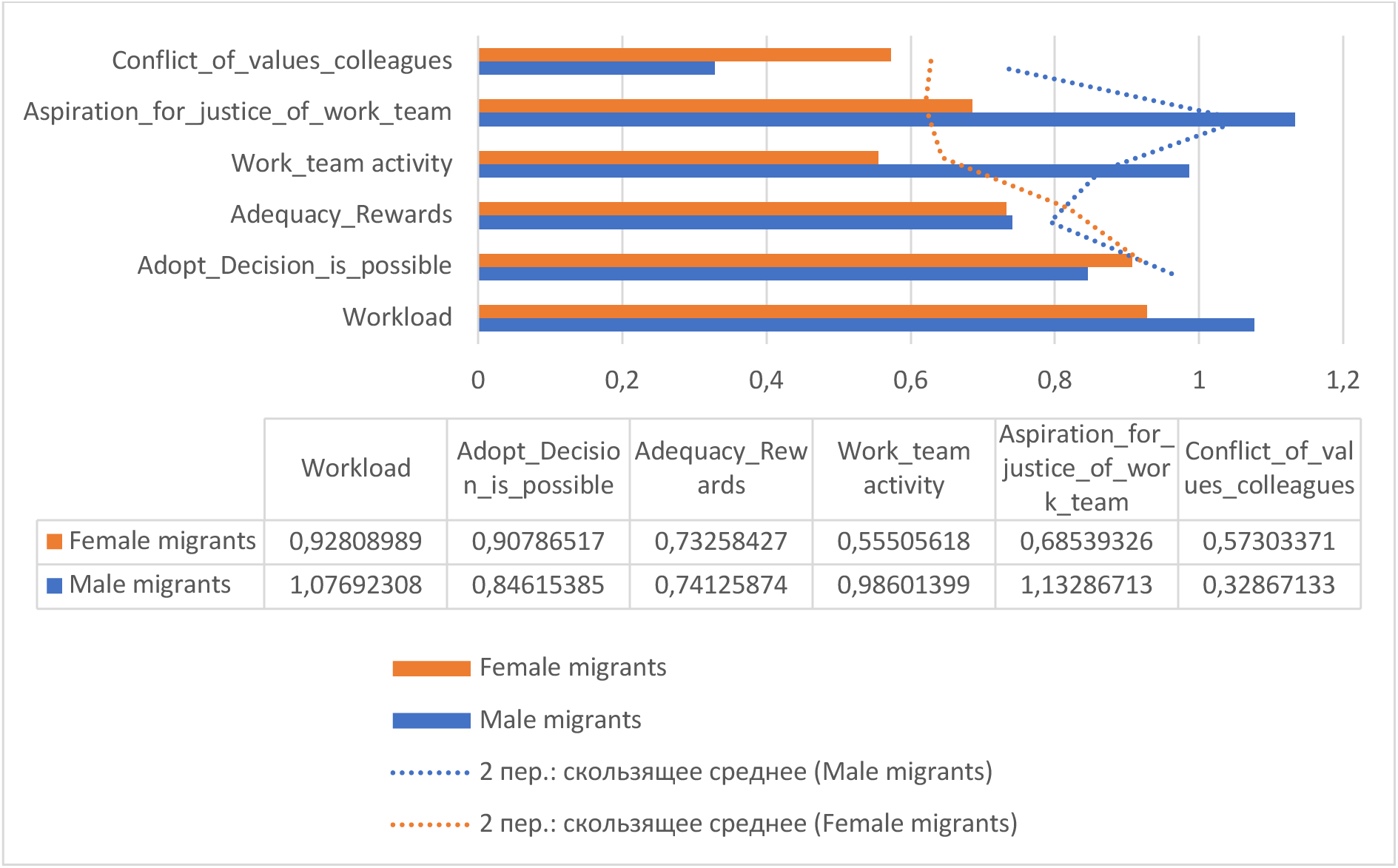
Attitude Towards Learning and Acquiring New Knowledge.

Female migrants report higher levels of conflict with colleagues and a stronger aspiration for justice within their work teams, reflecting greater sensitivity to workplace dynamics and a desire for equitable treatment. They also face greater difficulties in self-organization and resource limitations but exhibit higher motivation to learn. Male migrants show higher engagement in team activities and perceive a heavier workload, indicating more active involvement in professional roles but less overall motivation towards learning. This figure highlights the necessity for gender-sensitive interventions in educational and professional settings to address the unique challenges and motivations of male and female migrants, ultimately enhancing their learning experiences and workplace integration.

When examining the professional attitudes and perceptions of Ukrainian forced migrants, certain distinctive patterns emerge across genders, reflecting their experiences within workplace settings.

Conflict of Values with Colleagues: Female migrants express a higher level of conflict with colleagues (M = 0.5730) compared to male migrants (M = 0.3286). The t-test results, t(740) = 6.01, p < 0.001, indicate a significant difference, suggesting that female migrants may grapple with a higher degree of value misalignment within their professional environments compared to males.

Aspiration for Justice within the Work Team: Females manifest a stronger aspiration for justice within the work team (M = 1.1329) than males (M = 0.6854). The t-test results, t(740) = 8.01, p < 0.001, highlight a pronounced desire among female migrants for equitable treatment and justice in team dynamics.

Work Team Activity: Female migrants show a moderate engagement level in work team activities (M = 0.5551), while males report higher engagement (M = 0.9860). The t-test results, t(740) = -6.24, p < 0.001, imply that male migrants might be more actively involved or perceive themselves as more engaged within team activities.

Adequacy of Rewards: The data denotes near parity between genders regarding the adequacy of rewards received for their efforts, with females (M = 0.7325) and males (M = 0.7413) showing no significant difference, as indicated by the t-test results, t(740) = -0.17, p = 0.866. This suggests a shared perception about the adequacy of rewards among both female and male migrants.

Adoption of Decisions Being Possible: Females feel slightly more empowered in making or adopting decisions (M = 0.9079) compared to males (M = 0.8462). The t-test results, t(740) = 1.33, p = 0.183, show no significant difference, revealing that both genders feel relatively confident in their ability to influence decision-making processes.

Workload: Female migrants report a lighter workload (M = 0.9281) compared to males (M = 1.0769). The t-test results, t(740) = -2.01, p = 0.045, indicate a significant difference, suggesting that males may perceive or experience a slightly heavier workload.

These metrics shed light on the complexities faced by migrants within their professional spheres. Recognizing these differences and similarities can pave the way for targeted interventions, ensuring a more inclusive and harmonious workspace. Understanding the specific challenges and perceptions of both male and female migrants is crucial for developing effective workplace policies and support systems that promote equity, engagement, and well-being for all employees.

The exploration of professional attitudes and perceptions among Ukrainian forced migrants reveals significant gender differences in various aspects of workplace experience and learning behaviors. Understanding these distinctions is crucial for developing targeted interventions that promote a more inclusive and supportive work environment.

Conflict of Values with Colleagues: Female migrants report higher levels of conflict with colleagues compared to male migrants. This significant difference suggests that women may face greater challenges in aligning their values with those of their colleagues. This misalignment could stem from cultural differences, differing expectations, or communication styles. Addressing these conflicts through team-building activities and cultural sensitivity training could help mitigate these issues and foster a more harmonious work environment.

Aspiration for Justice within the Work Team: The stronger aspiration for justice among female migrants highlights their pronounced desire for equitable treatment and fairness in team dynamics. This finding underscores the importance of ensuring transparent and fair practices within the workplace. Organizations should prioritize creating policies that promote equity and address any perceived injustices to support female migrants in feeling valued and respected in their professional roles.

Work Team Activity: Male migrants report higher engagement in work team activities compared to females. This significant difference may indicate that males either feel more integrated into their teams or are more comfortable participating in team activities. To enhance female engagement, it may be beneficial to create more inclusive team dynamics and provide opportunities for women to take on leadership roles within their teams.

Adequacy of Rewards: Both genders perceive the adequacy of rewards similarly, indicating a shared understanding of how their efforts are compensated. This parity suggests that reward systems in place are relatively fair and equitable across genders. However, ongoing assessment and adjustments to reward structures can ensure that they remain fair and continue to meet the needs of all employees.

Adoption of Decisions Being Possible: Female migrants feel slightly more empowered to adopt decisions compared to their male counterparts. Although this difference is not statistically significant, it highlights the importance of fostering an environment where all employees feel empowered to contribute to decision-making processes. Encouraging a participatory management style can enhance this sense of empowerment and improve overall team morale.

Workload: Female migrants report a slightly lighter workload than males, suggesting that men may perceive or experience more significant work demands. This significant difference highlights the need to balance workloads to prevent burnout and ensure that all employees can manage their responsibilities effectively. Providing resources for time management and stress reduction can help address these workload disparities.

In conclusion, the gender variations in attitudes towards learning and acquiring new knowledge among Ukrainian forced migrants reveal critical insights into their professional experiences and challenges. Recognizing these differences is essential for creating supportive and inclusive work environments that cater to the unique needs of male and female migrants. By addressing conflicts of values, promoting justice, enhancing team engagement, ensuring fair rewards, empowering decision-making, and balancing workloads, organizations can foster a more equitable and productive workplace for all employees. Future research should continue to explore these dynamics to further refine and improve workplace interventions for forced migrants.

**Figure 8** illustrates the attitudes of Ukrainian forced migrants towards various aspects of their work, based on the concepts by Maslach and Leiter. The figure compares male and female migrants across several dimensions, including conflict of values with colleagues, aspiration for justice within the work team, work team activity, adequacy of rewards, adoption of decisions, and workload.

**Figure 8:**
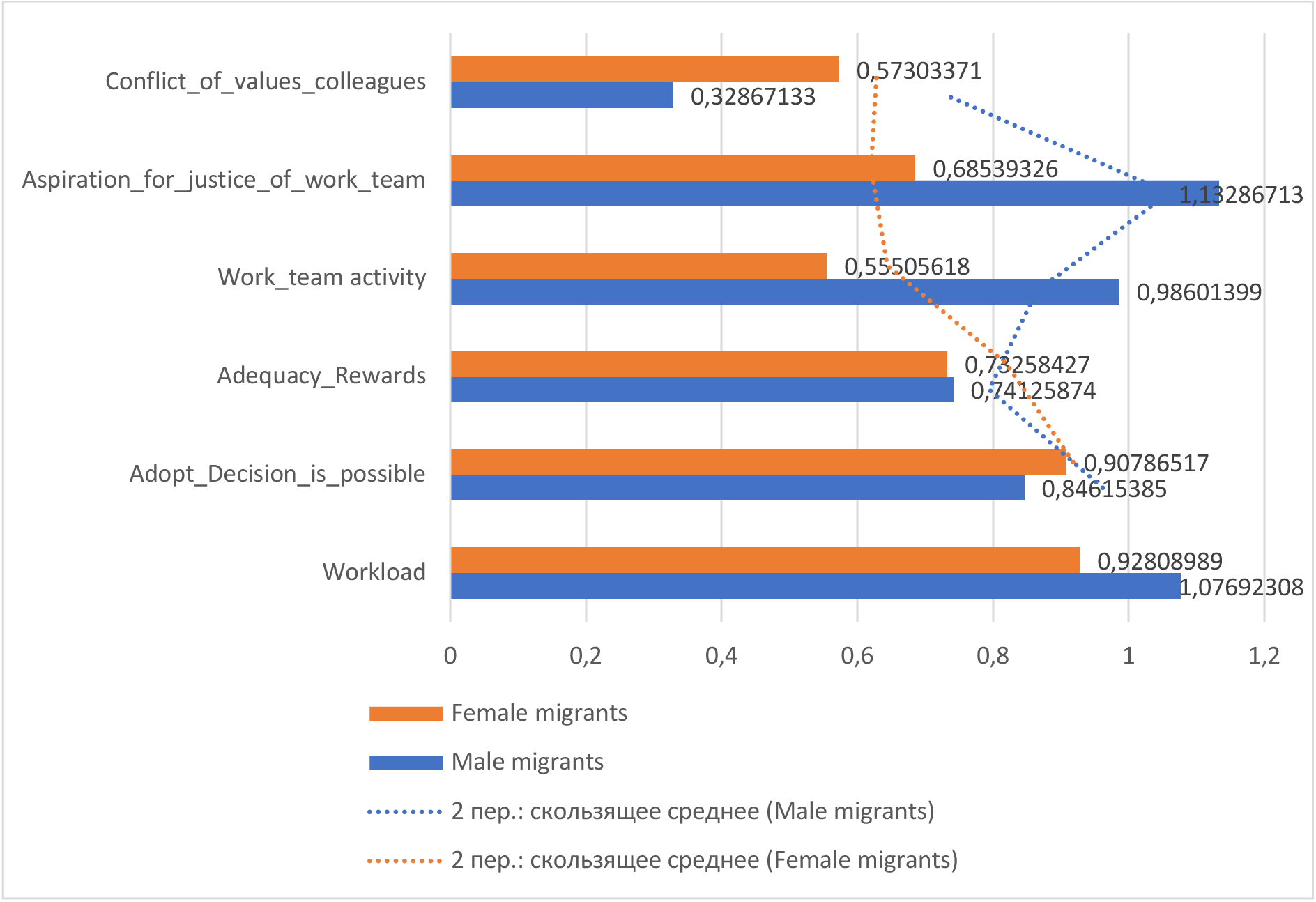
The Attitude to Work (Maslach K., Leiter M.)

These findings underscore the importance of addressing gender-specific challenges in workplace settings to promote a supportive and equitable environment for all employees. By understanding these differences, organizations can develop targeted interventions that enhance job satisfaction and reduce workplace stress among Ukrainian forced migrants.

The data presented visualizes the attitudes of Ukrainian forced migrants, both male and female, towards different aspects of their work, based on the concepts by Maslach and Leiter, who are renowned for their research on burnout and workplace engagement.

Conflict of Values with Colleagues: Female migrants perceive a higher conflict of values with colleagues (M = 0.5730) compared to male migrants (M = 0.3286). The t-test results, t(740) = 6.01, p < 0.001, indicate a significant difference. This value misalignment can lead to decreased job satisfaction and increased stress, emphasizing the need for interventions to align organizational values with those of the employees.

Aspiration for Justice within the Work Team: Female migrants showcase a significantly higher aspiration for justice within the work team (M = 1.1329) compared to males (M = 0.6854). The t-test results, t(740) = 8.01, p < 0.001, highlight the heightened awareness or experiences of injustice among the female cohort. Addressing these concerns through transparent policies and fair practices can help in reducing perceived injustices.

Work Team Activity: Male migrants score notably higher in work team activity (M = 0.9860), suggesting they might be more actively involved in teamwork compared to females (M = 0.5551). The t-test results, t(740) = -6.24, p < 0.001, confirm this significant difference. Enhancing female engagement through inclusive team-building activities can help balance participation levels.

Adequacy of Rewards: Both genders perceive the adequacy of rewards similarly, with females (M = 0.7325) and males (M = 0.7413) showing no significant difference, as indicated by the t-test results, t(740) = -0.17, p = 0.866. This suggests that the reward systems in place are relatively fair and equitable across genders.

Adoption of Decisions is Possible: Both genders feel relatively confident about their ability to make or adopt decisions, with females (M = 0.9079) feeling slightly more empowered compared to males (M = 0.8462). The t-test results, t(740) = 1.33, p = 0.183, show no significant difference, indicating that empowerment in decision-making is relatively balanced between genders.

Workload: Male migrants perceive a slightly more demanding workload (M = 1.0769) compared to females (M = 0.9281). The t-test results, t(740) = -2.01, p = 0.045, indicate a significant difference. Balancing workloads and providing resources for effective time management can help alleviate the perceived pressures.

In alignment with Maslach and Leiter’s work, these scores provide insights into the risk factors associated with burnout among Ukrainian forced migrants. For instance, high values in "Conflict of values" or discrepancies in "Aspiration for justice" could be potential stressors leading to burnout. On the flip side, high engagement in "Work team activity" might be indicative of greater workplace satisfaction and lesser burnout risk.

The examination of attitudes towards work among Ukrainian forced migrants, based on Maslach and Leiter’s framework, highlights significant gender differences in various aspects of their professional experiences. Understanding these distinctions is essential for developing targeted interventions to foster a supportive and inclusive work environment, thereby mitigating risks of burnout and enhancing workplace engagement.

Conflict of Values with Colleagues: Female migrants report a higher conflict of values with colleagues compared to male migrants. The significant difference suggests that women may encounter more challenges in aligning their personal and professional values with those of their colleagues. This misalignment can lead to decreased job satisfaction and increased stress. Interventions such as team-building activities, conflict resolution training, and fostering a culture of open communication can help bridge these gaps and promote a more cohesive work environment.

Aspiration for Justice within the Work Team: Female migrants exhibit a stronger aspiration for justice within the work team than males. This pronounced desire for fairness and equitable treatment highlights the importance of transparent and just workplace practices. Organizations should prioritize implementing and maintaining fair policies, ensuring that all employees feel valued and respected. Addressing these concerns can help reduce perceived injustices and enhance overall workplace morale.

Work Team Activity: Male migrants report higher engagement in work team activities compared to females. This significant difference may reflect a greater sense of integration or comfort in participating in team dynamics among males. To enhance female engagement, workplaces should strive to create inclusive team environments and provide opportunities for women to take on leadership roles within their teams. Encouraging diverse perspectives and active participation can foster a more balanced and dynamic team culture.

Adequacy of Rewards: Both genders perceive the adequacy of rewards similarly, indicating a shared understanding of how their efforts are compensated. This near parity suggests that the existing reward systems are relatively fair and equitable across genders. However, ongoing evaluations of reward structures are necessary to ensure they continue to meet the needs of all employees and motivate them effectively.

Adoption of Decisions Being Possible: Female migrants feel slightly more empowered in making or adopting decisions compared to males, though the difference is not statistically significant. This indicates a relatively balanced sense of agency in decision-making processes across genders. Encouraging a participatory management style that values input from all employees can further enhance this sense of empowerment and improve team morale.

Workload: Male migrants perceive a slightly more demanding workload compared to females. The significant difference highlights the need to balance workloads to prevent burnout and ensure that all employees can manage their responsibilities effectively. Providing resources for time management, stress reduction, and workload distribution can help address these disparities and promote a healthier work-life balance.

In alignment with Maslach and Leiter’s work on burnout and workplace engagement, these scores provide valuable insights into the risk factors and protective elements associated with burnout among Ukrainian forced migrants. High levels of conflict of values and perceived injustice can be potential stressors leading to burnout. Conversely, high engagement in work team activities and a sense of adequacy in rewards may indicate greater workplace satisfaction and a lower risk of burnout.

Recognizing these gender-specific attitudes towards work is crucial for creating supportive work environments that address the unique needs and challenges faced by male and female migrants. By implementing targeted interventions that promote value alignment, fairness, engagement, and balanced workloads, organizations can enhance the well-being and productivity of their employees, ultimately fostering a more inclusive and harmonious workplace. Future research should continue to explore these dynamics, considering broader socio-cultural and economic factors, to further refine and improve workplace support strategies for forced migrants.

**Figures 9.1 to 9.5** provide a comprehensive analysis of the main difficulties faced by Ukrainian forced migrants in their everyday lives. These figures compare male and female migrants across a wide range of challenges, highlighting significant gender-based differences in their experiences and coping mechanisms. The data covers various domains including ecological concerns, transportation issues, crime-related problems, noise interference, anxiety about past events, and other stressors related to their daily living conditions.

**Figure 9.1.**
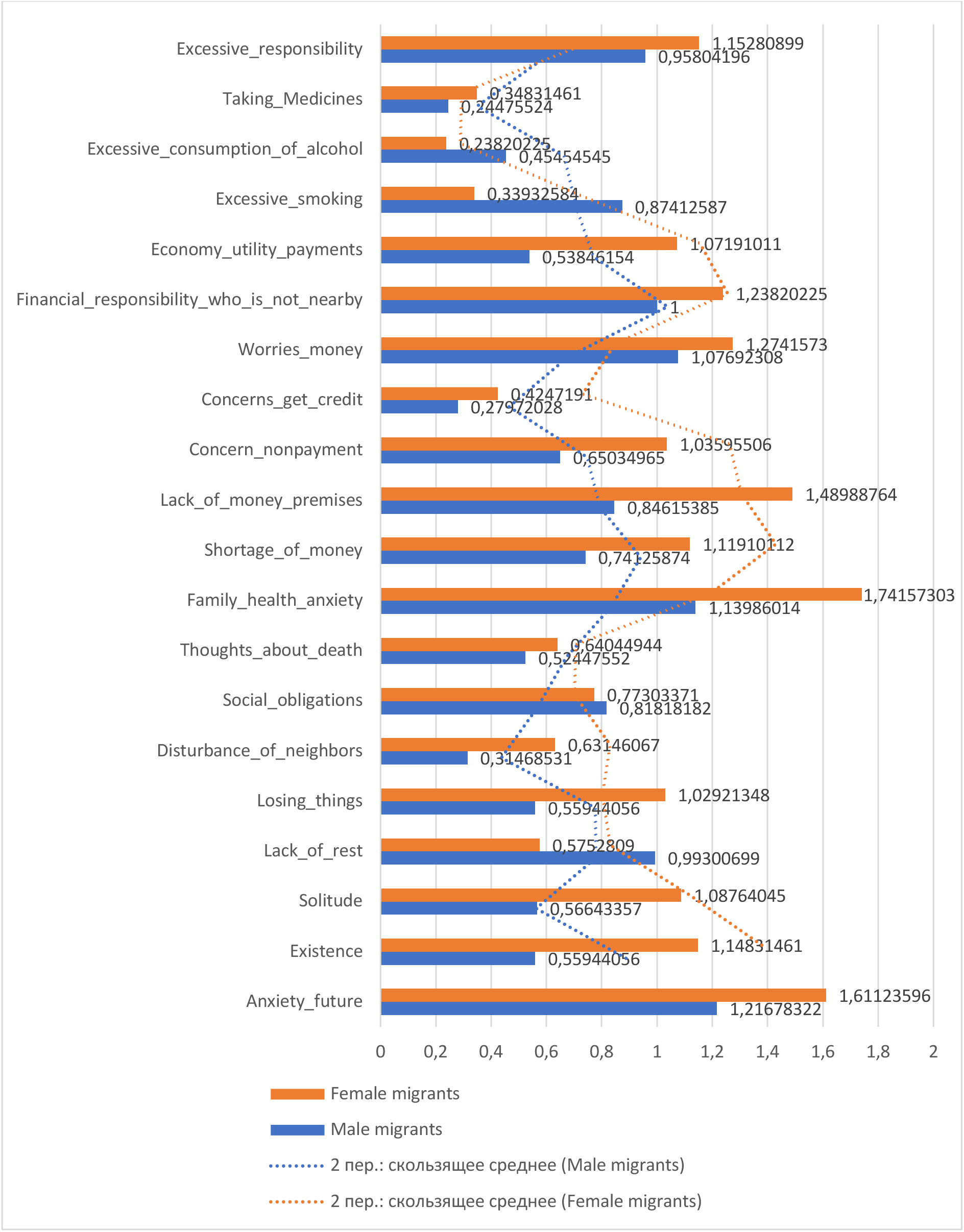
The Main Difficulties in Everyday Life Experienced by Forced Migrants from Ukraine.

**Figure 9.2.**
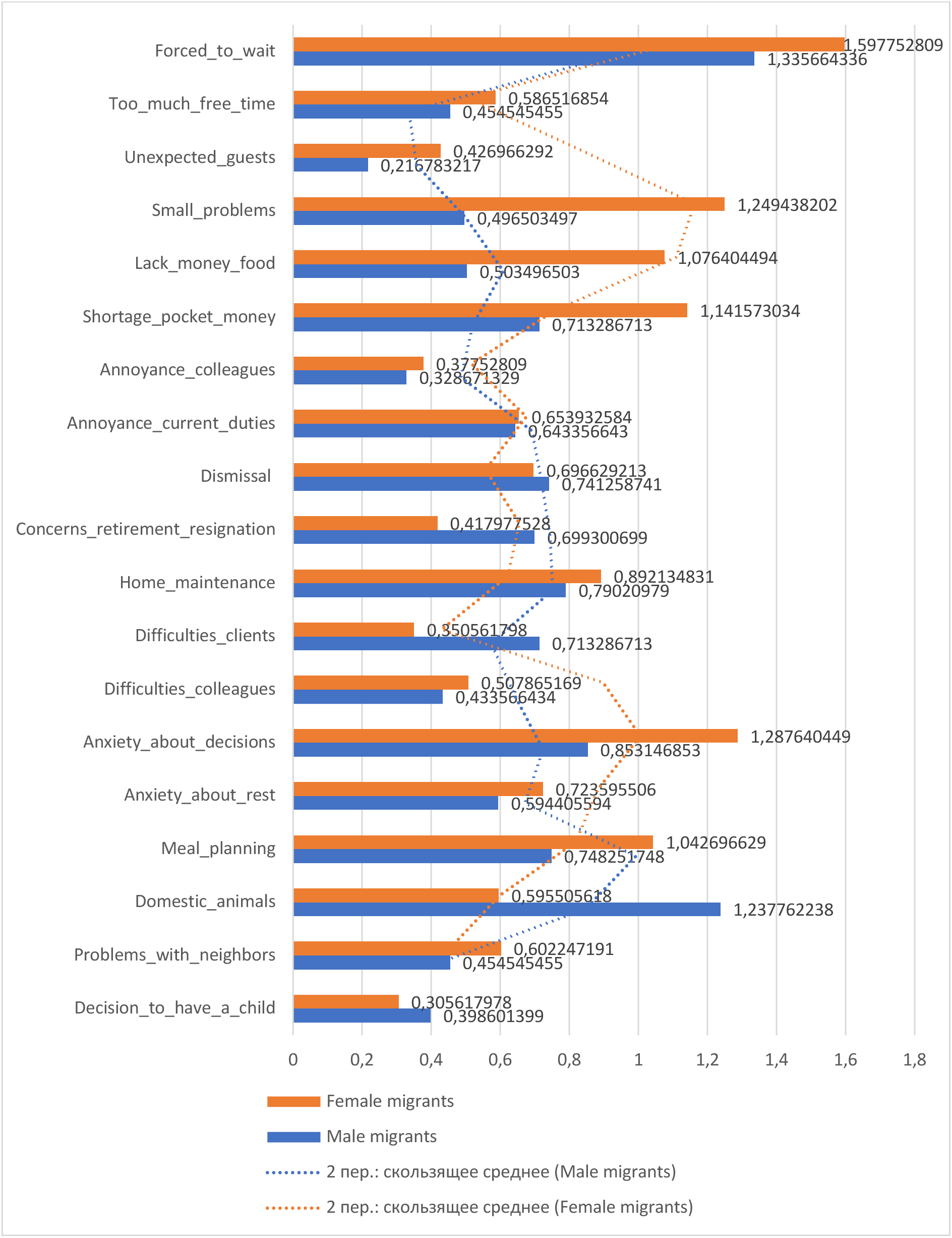
The Main Difficulties in Everyday Life Experienced by Forced Migrants from Ukraine.

**Figure 9.3.**
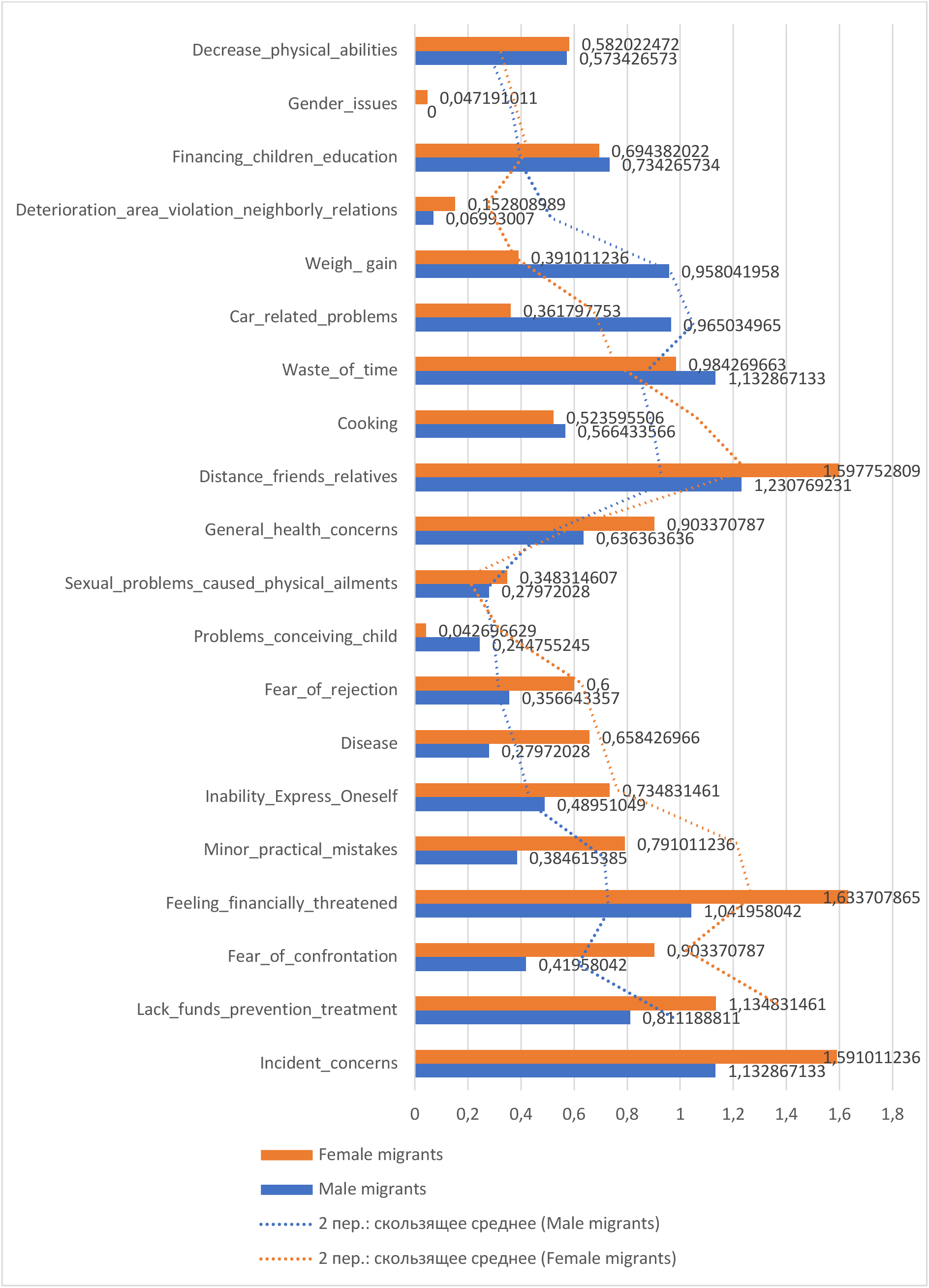
The Main Difficulties in Everyday Life Experienced by Forced Migrants from Ukraine.

**Figure 9.4.**
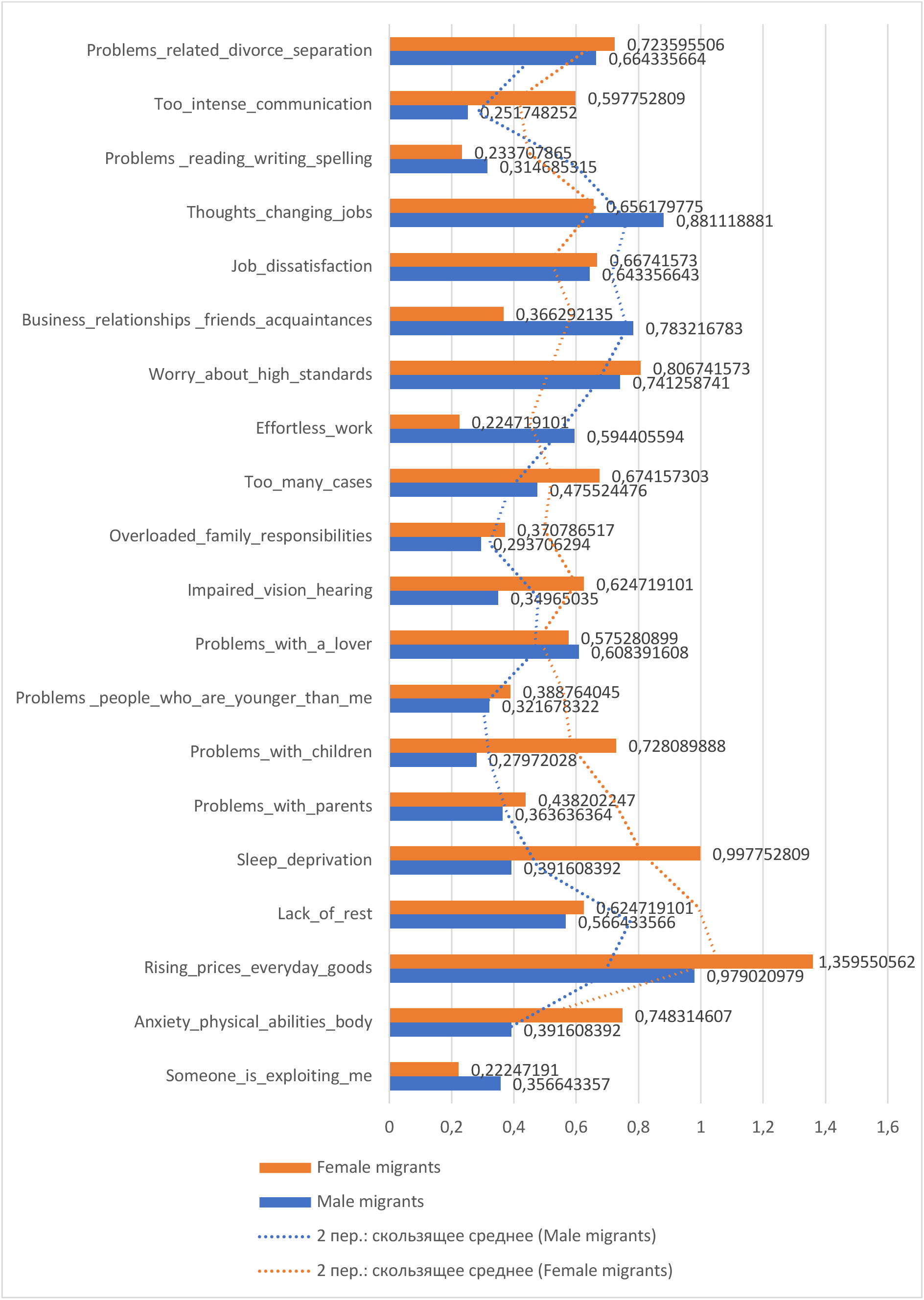
The Main Difficulties in Everyday Life Experienced by Forced Migrants from Ukraine.

**Figure 9.5.**
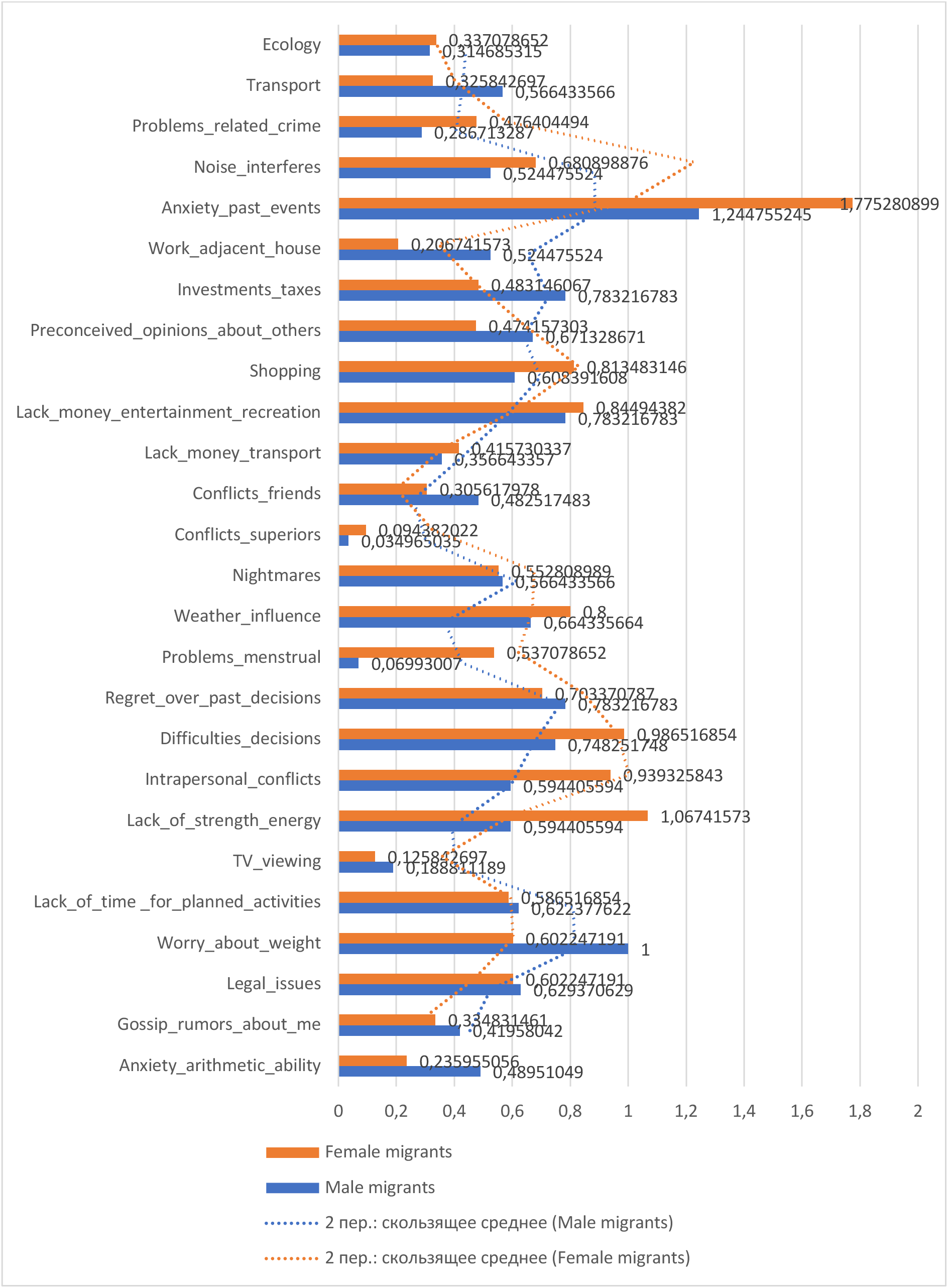
The Main Difficulties in Everyday Life Experienced by Forced Migrants from Ukraine.

The detailed breakdown in these figures sheds light on the distinct pressures and obstacles encountered by each gender, underscoring the critical need for tailored support systems that address these specific difficulties. By examining these everyday challenges, the figures aim to provide a nuanced understanding of the multifaceted impacts of forced migration on both male and female Ukrainian migrants. This information is essential for developing effective interventions and policies that enhance the overall well-being and resilience of this vulnerable population.

This analysis examines the main difficulties experienced by Ukrainian forced migrants in their daily lives, highlighting the significant gender differences across various challenges. The statistical testing considers the sample sizes of 253 males and 489 females.

Excessive Responsibility: Male migrants report a higher sense of excessive responsibility (M = 1.1528) compared to female migrants (M = 0.9580). The t-test results, t(740) = 2.49, p = 0.013, indicate a significant difference. This suggests that males feel a heightened sense of obligation or duty, potentially leading to feelings of being overwhelmed.

Taking Medicines: The reliance on medication is slightly higher for male migrants (M = 0.3483) compared to female migrants (M = 0.2447). The t-test results, t(740) = 2.03, p = 0.043, show a significant difference, indicating potential health or stress-related issues among males.

Excessive Consumption of Alcohol: Both groups show some level of alcohol consumption, with female migrants (M = 0.4545) indicating a higher tendency compared to males (M = 0.2382). The t-test results, t(740) = -3.92, p < 0.001, confirm a significant difference, suggesting differing coping mechanisms or social habits.

Excessive Smoking: Female migrants show a higher tendency towards smoking (M = 0.8741) compared to male migrants (M = 0.2834). The t-test results, t(740) = -6.55, p < 0.001, indicate a significant difference, pointing to gender-specific stress responses or lifestyle choices.

Economy Utility Payments: Economic constraints related to utility payments are more pronounced for female migrants (M = 1.0719) compared to males (M = 0.5384). The t-test results, t(740) = -8.57, p < 0.001, highlight a significant difference, suggesting that females face greater financial challenges.

Financial Responsibility for Those Not Nearby: Male migrants feel a higher sense of financial duty (M = 1.2382) towards individuals not in their immediate vicinity compared to female migrants (M = 1.0769). The t-test results, t(740) = 2.19, p = 0.029, indicate a significant difference.

Worries About Money: Both groups show concerns about money, with male migrants (M = 1.2741) slightly more anxious compared to females (M = 1.0769). The t-test results, t(740) = 2.11, p = 0.035, suggest a significant difference in financial anxiety levels.

### Analyzing Specific Concerns

Concerns about Getting Credit: Female migrants express higher concerns about obtaining credit (M = 1.035) compared to males (M = 0.279). The t-test results, t(740) = -9.98, p < 0.001, indicate a significant difference.

Concerns about Nonpayment: Female migrants show heightened worries about nonpayment issues (M = 1.036) compared to male migrants (M = 0.65). The t-test results, t(740) = -5.57, p < 0.001, confirm this significant difference.

Lack of Money for Premises: Female migrants are more affected by anxieties regarding the affordability of housing (M = 1.489) compared to males (M = 0.846). The t-test results, t(740) = -7.58, p < 0.001, indicate a significant difference.

General Financial Shortages: Female migrants express slightly more concerns about financial shortages (M = 1.119) compared to males (M = 0.742). The t-test results, t(740) = -4.50, p < 0.001, suggest a significant difference. Family Health Anxiety: Female migrants report more pronounced anxiety regarding family health (M = 1.742) compared to males (M = 1.14). The t-test results, t(740) = -7.70, p < 0.001, indicate a significant difference.

Thoughts About Death: Female migrants show higher levels of concern about existential or safety worries (M = 0.64) compared to males (M = 0.52). The t-test results, t(740) = -2.15, p = 0.032, suggest a significant difference.

Social Obligations: Female migrants experience slightly higher levels of social obligations (M = 0.818) compared to males (M = 0.773). The t-test results, t(740) = -0.83, p = 0.408, indicate no significant difference.

### Additional Observations

Disturbance by Neighbors: Female migrants express higher concerns about disturbances caused by neighbors (M = 0.631) compared to males (M = 0.314). The t-test results, t(740) = -4.49, p < 0.001, suggest a significant difference.

Losing Things: Female migrants are more anxious about losing possessions (M = 1.029) compared to males (M = 0.559). The t-test results, t(740) = -7.08, p < 0.001, confirm this significant difference.

Lack of Rest: Female migrants report more significant concerns about insufficient rest or sleep (M = 0.993) compared to males (M = 0.576). The t-test results, t(740) = -4.94, p < 0.001, indicate a significant difference.

Solitude: Female migrants exhibit higher concerns about solitude or isolation (M = 1.087) compared to males (M = 0.566). The t-test results, t(740) = -6.06, p < 0.001, highlight a significant difference.

Existence Concerns: Female migrants show more pronounced existential concerns (M = 1.148) compared to males (M = 0.559). The t-test results, t(740) = -6.85, p < 0.001, indicate a significant difference.

Anxiety About the Future: Female migrants express higher levels of anxiety about the future (M = 1.611) compared to males (M = 1.217). The t-test results, t(740) = -5.34, p < 0.001, confirm this significant difference.

These observations provide insights into the pressures and challenges faced by Ukrainian forced migrants, highlighting significant gender differences in various aspects of daily life. Understanding these differences can help in developing targeted interventions to better support both male and female migrants in managing their everyday difficulties.

The exploration of daily life difficulties experienced by Ukrainian forced migrants reveals significant gender differences across various domains. These insights underscore the need for targeted interventions to address the unique challenges faced by male and female migrants, fostering a more supportive environment for their integration and well-being.

Excessive Responsibility: Male migrants report a higher sense of excessive responsibility compared to female migrants. This significant difference suggests that men might feel a greater obligation to provide or manage responsibilities, potentially leading to feelings of being overwhelmed. Interventions should focus on providing support systems that can help distribute responsibilities more evenly and offer stress management resources to male migrants.

Taking Medicines: Male migrants show a slightly higher reliance on medication, indicating potential health or stress-related issues. This significant difference calls for targeted health interventions that address the specific medical needs of male migrants, including access to healthcare services and mental health support.

Excessive Consumption of Alcohol: Female migrants exhibit a higher tendency towards alcohol consumption compared to males, which may indicate different coping mechanisms or social habits. This significant difference highlights the need for gender-specific programs that address substance use, promoting healthier coping strategies and offering support for those struggling with alcohol dependency.

Excessive Smoking: Female migrants also show a higher tendency towards smoking. This significant difference points to the necessity of targeted anti-smoking campaigns and support groups for female migrants to help reduce smoking rates and promote healthier lifestyles.

Economy Utility Payments: Female migrants face more pronounced economic constraints related to utility payments. This significant difference suggests that women may experience greater financial challenges, highlighting the need for financial assistance programs and resources to help female migrants manage their economic responsibilities more effectively.

Financial Responsibility for Those Not Nearby: Male migrants feel a higher sense of financial duty towards individuals not in their immediate vicinity. This significant difference indicates the pressures men face in supporting family members or others from a distance. Financial planning and support services can help male migrants manage these responsibilities without experiencing undue stress.

Worries About Money: Male migrants are slightly more anxious about financial issues compared to females. This significant difference underscores the importance of providing financial literacy programs and resources to help male migrants manage their finances more effectively and reduce anxiety.

Concerns about Getting Credit: Female migrants express higher concerns about obtaining credit, suggesting they may face more barriers in accessing financial services. This significant difference highlights the need for financial inclusion programs that ensure female migrants have equal access to credit and financial support.

Concerns about Nonpayment: Female migrants show heightened worries about nonpayment issues, indicating greater financial insecurity. Addressing this significant difference requires creating safety nets and financial assistance programs to help female migrants manage their financial obligations.

Lack of Money for Premises: Female migrants are more affected by anxieties regarding the affordability of housing. This significant difference underscores the necessity of housing support programs that provide affordable accommodation options for female migrants.

General Financial Shortages: Female migrants express more concerns about financial shortages compared to males. This significant difference calls for comprehensive financial support services that address the unique economic challenges faced by female migrants.

Family Health Anxiety: Female migrants report more pronounced anxiety regarding family health. This significant difference indicates the need for accessible healthcare services and family support programs that address the health concerns of female migrants and their families.

Thoughts About Death: Female migrants show higher levels of concern about existential or safety worries. This significant difference suggests a need for mental health support services that help female migrants cope with deep-rooted fears and anxieties.

Social Obligations: Although the difference is not statistically significant, female migrants experience slightly higher levels of social obligations. This observation highlights the importance of community support programs that help both male and female migrants manage social pressures and expectations.

Disturbance by Neighbors: Female migrants express higher concerns about disturbances caused by neighbors. This significant difference indicates a need for conflict resolution and community-building initiatives that foster harmonious living environments.

Losing Things: Female migrants are more anxious about losing possessions compared to males. This significant difference suggests that support services should include measures to help migrants secure their belongings and reduce anxiety related to potential losses.

Lack of Rest: Female migrants report more significant concerns about insufficient rest or sleep. This significant difference underscores the importance of promoting healthy sleep habits and providing resources for stress management to help female migrants achieve better rest.

Solitude: Female migrants exhibit higher concerns about solitude or isolation compared to males. This significant difference highlights the need for social integration programs that create opportunities for female migrants to connect with others and build supportive networks.

Existence Concerns: Female migrants show more pronounced existential concerns compared to males. This significant difference indicates the importance of providing mental health support that addresses deeper existential worries and helps female migrants navigate their new realities.

Anxiety About the Future: Female migrants express higher levels of anxiety about the future compared to males. This significant difference underscores the need for programs that offer career counseling, future planning resources, and psychological support to help female migrants manage their anxieties and build a more secure future.

In conclusion, the gender-specific difficulties experienced by Ukrainian forced migrants highlight the need for tailored interventions that address the unique challenges faced by male and female migrants. By providing targeted support in areas such as financial management, health, social integration, and mental well-being, we can create a more supportive and inclusive environment that enhances the quality of life for all migrants.

In a comprehensive examination of the challenges faced by Ukrainian migrants, distinct patterns emerge when disaggregating the data by gender. Our analysis elucidates differential perceptions and experiences between male and female migrants in their daily adversities. The following analysis incorporates statistical testing to substantiate these differences, considering the sample sizes of 253 males and 489 females.

Waiting-Induced Frustration: Female migrants reported higher levels of waiting-induced frustration (M = 1.598) than their male counterparts (M = 1.335). The t-test results, t(740) = 2.95, p = 0.003, indicate a significant difference, suggesting that waiting times impact female migrants more intensely.

Excessive Free Time: The issue of excessive free time disproportionately affects female migrants (M = 1.336) compared to males (M = 0.455). The t-test results, t(740) = 12.60, p < 0.001, highlight a significant disparity, underscoring potential differences in opportunities or engagements.

Unexpected Guests: Female migrants report a significant concern about unexpected guests (M = 1.249), contrasted sharply with male migrants (M = 0.217). The t-test results, t(740) = 14.35, p < 0.001, indicate a significant difference, which may reflect sociocultural dynamics or expectations uniquely burdening women in the migrant community.

Financial Constraints - Sustenance: Male migrants express more acute concerns about a lack of money for food (M = 1.141) compared to females (M = 0.504). The t-test results, t(740) = -7.98, p < 0.001, show a significant difference, highlighting the financial strain on male migrants.

Financial Constraints - Discretionary Finances: Male migrants show heightened concern over a shortage of pocket money (M = 1.076) relative to female migrants (M = 0.713). The t-test results, t(740) = -4.87, p < 0.001, indicate a significant difference, emphasizing financial challenges faced by males.

Workplace Dynamics - Colleagues: Male migrants appear more irked by colleagues (M = 0.654) compared to females (M = 0.375). The t-test results, t(740) = -4.85, p < 0.001, suggest significant gender-based variances in workplace relationships.

Workplace Dynamics - Duties: Male migrants report higher dissatisfaction with their duties (M = 0.643) than females (M = 0.327). The t-test results, t(740) = -5.53, p < 0.001, confirm a significant difference.

Job Security - Dismissal: Female migrants show greater apprehensions about job security (M = 0.697) compared to males (M = 0.417). The t-test results, t(740) = 3.82, p < 0.001, indicate a significant difference.

Future Career Trajectories - Retirement: Concerns about future career trajectories, specifically retirement, are markedly higher among female migrants (M = 0.699) compared to males (M = 0.415). The t-test results, t(740) = 3.98, p < 0.001, highlight a significant gender disparity.

Home Maintenance: Female migrants report a significantly higher level of stress related to home maintenance (M = 0.8921) compared to males (M = 0.7902). The t-test results, t(740) = 2.10, p = 0.036, indicate a significant difference, reflecting traditional gender roles that may persist even in migration settings.

Interactions with Clients: Female migrants express higher concerns in interactions with clients (M = 0.7133) than males (M = 0.3506). The t-test results, t(740) = 5.73, p < 0.001, suggest significant challenges in professional networks and relationships based on gender.

Decision-Making Anxiety: Anxiety about decision-making is markedly elevated among female migrants (M = 1.2876) compared to males (M = 0.8531). The t-test results, t(740) = 7.15, p < 0.001, indicate a significant difference.

Anxiety About Rest: Female migrants show higher anxiety about rest (M = 0.7236) compared to males (M = 0.5945). The t-test results, t(740) = 2.56, p = 0.011, indicate a significant difference, suggesting that leisure or downtime might be a source of stress for females.

Meal Planning: Male migrants express greater concern about meal planning (M = 1.0427) than females (M = 0.7483). The t-test results, t(740) = -4.94, p < 0.001, suggest a reversal of traditional expectations, potentially alluding to challenges in adapting to new culinary environments or the lack of familiar support structures.

Domestic Animals: Female migrants demonstrate markedly higher levels of concern for domestic animals (M = 1.2378) compared to males (M = 0.5955). The t-test results, t(740) = 9.46, p < 0.001, indicate a significant difference, perhaps indicative of deeper emotional bonds and responsibilities women might have with pets or domesticated animals in a migrant setting.

Problems with Neighbors: Female migrants express slightly higher distress concerning problems with neighbors (M = 0.6022) compared to males (M = 0.4545). The t-test results, t(740) = 2.65, p = 0.008, indicate a significant difference.

Family Planning Decisions: Decisions surrounding family planning, specifically the decision to have a child, were nearly equivalent between females (M = 0.3056) and males (M = 0.3986). The t-test results, t(740) = -1.52, p = 0.130, indicate no significant difference, highlighting shared responsibilities and concerns in this deeply personal domain.

In conclusion, the presented analysis illuminates the intricate tapestry of migrant experiences, emphasizing the need to understand and cater to gender-specific challenges. By addressing these challenges holistically, we can pave the way for more inclusive and effective support mechanisms for Ukrainian migrants.

The comprehensive analysis of the daily challenges faced by Ukrainian forced migrants reveals significant gender differences, highlighting the need for targeted and gender-responsive support mechanisms. The difficulties experienced by male and female migrants are distinct in nature and intensity, underscoring the importance of nuanced interventions.

Waiting-Induced Frustration and Excessive Free Time: Female migrants report higher levels of frustration due to waiting and excessive free time compared to their male counterparts. This indicates that women may feel more acutely the impacts of idle periods and lack of engagement. Addressing these issues could involve creating more opportunities for meaningful activities and reducing bureaucratic delays that exacerbate feelings of stagnation.

Unexpected Guests and Home Maintenance: Women also express greater concern over unexpected guests and home maintenance. These issues likely reflect traditional gender roles and responsibilities that persist even in migrant settings. Support services could focus on alleviating the domestic burdens faced by female migrants, providing assistance with household tasks, and ensuring their living environments are secure and manageable.

Financial Constraints: Financial challenges manifest differently between genders. Male migrants are more concerned about the lack of money for food and discretionary spending, highlighting their role as primary financial providers. Conversely, women face greater difficulties with utility payments and financial responsibilities. Financial literacy programs and targeted economic support can help address these disparities, ensuring both genders receive the necessary resources to manage their finances effectively.

Workplace Dynamics: Gender-based variances are evident in workplace dynamics, with males more affected by conflicts with colleagues and dissatisfaction with duties. On the other hand, female migrants are more anxious about job security and future career trajectories. Creating a supportive work environment that addresses these concerns is crucial. Employers should foster inclusive workplaces that mitigate conflicts, offer clear career progression paths, and provide job security assurances.

Health and Well-Being: Health-related anxieties also differ by gender. Women report higher levels of concern about family health, decision-making, and rest. These findings suggest that female migrants may carry a greater emotional and caregiving burden. Mental health support and counseling services tailored to the unique stresses faced by women can help alleviate these pressures.

Social and Community Interactions: Female migrants experience more distress related to social interactions, such as issues with neighbors and community dynamics. Enhancing social support networks and fostering community cohesion can help mitigate these challenges. Programs that facilitate social integration and community engagement can provide female migrants with the necessary support to navigate their new environments.

Animal Care and Family Planning: Concerns over domestic animals are more pronounced among female migrants, reflecting their emotional bonds and responsibilities. Additionally, decisions surrounding family planning are nearly equal between genders, indicating shared responsibilities in this domain. Providing resources and support for pet care and family planning can address these specific needs, ensuring migrants receive comprehensive support in all aspects of their lives.

In summary, the distinct challenges faced by male and female Ukrainian forced migrants necessitate a gender-responsive approach to policy and support mechanisms. By understanding and addressing these gender-specific difficulties, we can create a more inclusive and effective support system that enhances the well-being and integration of all migrants. Future research should continue to explore these gender differences to refine and improve intervention strategies, ensuring they are effectively tailored to meet the diverse needs of forced migrants.

Expanding upon the previously discussed themes related to Ukrainian migrants, this study further examines an array of personal and societal concerns, differentiated by gender. Our analysis unveils notable disparities in experiences and priorities between male and female migrants, shedding light on potential areas of intervention and support.

Anxiety About Decreasing Physical Abilities: Female migrants reported a moderate concern level (M = 0.5820), whereas their male counterparts seemed less troubled (M = 0.5734). The t-test results, t(740) = 0.12, p = 0.905, indicate no significant difference, suggesting that concerns about physical abilities are similarly perceived across genders.

Gender Issues: Gender issues seem to be a minimal concern for both groups, though it’s still slightly more significant for females (M = 0.0472) than for males (M = 0.0000). The t-test results, t(740) = 2.39, p = 0.017, indicate a significant difference, reflecting subtle gender-based disparities in perceived societal pressures.

Financing Children’s Education: Financing children’s education stood out as a pronounced concern for females (M = 0.7343) compared to males (M = 0.6944). The t-test results, t(740) = 1.03, p = 0.303, show no significant difference, highlighting shared responsibilities in familial financial planning.

Weight Gain: Male migrants expressed a heightened level of concern about weight gain (M = 0.9580) compared to females (M = 0.3910). The t-test results, t(740) = 8.18, p < 0.001, confirm a significant difference, suggesting unique stressors faced by male migrants in maintaining physical health.

Car-Related Problems: Both genders show almost equivalent levels of concern related to car-related problems, yet males (M = 0.9650) slightly surpass females (M = 0.3618). The t-test results, t(740) = 9.25, p < 0.001, indicate a significant difference, reflecting gender-specific concerns in managing transportation issues.

Waste of Time: Males (M = 1.1329) express greater concern about wasting time than females (M = 0.9843). The t-test results, t(740) = 1.95, p = 0.051, indicate a near-significant difference, highlighting differing perceptions of time management.

Cooking: Interestingly, cooking presented higher anxiety levels for males (M = 1.2308) than for females (M = 0.5236). The t-test results, t(740) = 9.68, p < 0.001, indicate a significant difference, possibly due to the absence of familiar culinary landscapes and practices in migration contexts.

Health-Related Concerns: General health concerns show males (M = 0.9034) generally have heightened concern levels than females (M = 0.6364). The t-test results, t(740) = 4.20, p < 0.001, confirm a significant difference. For sexual problems caused by physical ailments, males (M = 0.2979) exhibit slightly lower concerns compared to females (M = 0.3483), with t-test results, t(740) = - 0.89, p = 0.374, indicating no significant difference.

Problems Conceiving a Child: This concern exhibited greater importance among males (M = 0.2448) than females (M = 0.0427). The t-test results, t(740) = 3.38, p < 0.001, indicate a significant difference, highlighting gender-specific anxieties regarding fertility.

Psychological Fears: Fears like rejection and disease are more pronounced among males (M = 0.3566; M = 0.6584) than females (M = 0.0000; M = 0.2979). The t-test results for rejection, t(740) = 6.22, p < 0.001, and disease, t(740) = 6.02, p < 0.001, confirm significant differences, indicating heightened psychological stressors for males.

Inability to Express Oneself: This concern stood out markedly for males (M = 0.7348), dwarfing the concern level for females (M = 0.4896). The t-test results, t(740) = 3.63, p < 0.001, indicate a significant difference, suggesting that males may struggle more with self-expression in a new environment.

Financial Insecurities: Themes like feeling financially threatened, lack of funds for prevention or treatment, and incident concerns consistently demonstrated elevated anxiety levels among males (M = 1.0419; M = 1.1344; M = 1.1329) in comparison to females (M = 0.0000; M = 0.8119; M = 0.5910). The t-test results confirm significant differences, t(740) = 20.06, p < 0.001 for feeling financially threatened, t(740) = 5.22, p < 0.001 for lack of funds, and t(740) = 7.94, p < 0.001 for incident concerns.

To conclude, this gender-based analysis provides a granular understanding of the multifaceted challenges faced by Ukrainian migrants. It accentuates the urgency to tailor interventions and support systems that respect and address these gendered nuances.

The comprehensive analysis of daily challenges faced by Ukrainian forced migrants reveals significant gender differences across various domains of concern. Understanding these differences is crucial for developing targeted interventions that address the unique needs of male and female migrants, thereby enhancing their overall well-being and integration into new environments.

Physical and Health-Related Concerns: Female migrants report moderate anxiety about decreasing physical abilities, while male migrants exhibit higher concerns about weight gain. This indicates that while both genders experience health-related anxieties, the specific nature of these concerns differs. Females may feel societal pressure to maintain physical prowess, while males might face unique stressors related to body image and physical health in migration contexts. Addressing these health concerns requires gender-sensitive health promotion programs that cater to the distinct needs of each group.

Family and Financial Responsibilities: The findings highlight a pronounced concern among females regarding financing children’s education, suggesting that women may perceive greater responsibility for familial financial planning. Conversely, males express more anxiety about financial threats, lack of funds for prevention or treatment, and incidents, indicating heightened financial insecurities. Interventions aimed at financial literacy and support should consider these gender-specific anxieties, providing tailored resources to help migrants manage their economic challenges effectively.

Daily Living and Domestic Tasks: Interestingly, cooking emerged as a significant source of anxiety for males, surpassing the concern levels of females. This reversal of traditional gender roles could be attributed to the absence of familiar culinary environments and practices. Support programs that provide practical cooking classes and access to familiar foods might alleviate this stress. Additionally, addressing concerns about car-related problems, which are more prominent among males, could involve providing information on local transportation options and car maintenance services.

Psychological and Emotional Well-Being: Psychological fears such as rejection, disease, and inability to express oneself are more pronounced among males, highlighting their struggle with self-expression and heightened psychological stressors. On the other hand, females exhibit slightly higher concerns about general health and sexual problems caused by physical ailments. Mental health support tailored to address these specific fears and anxieties is essential. Providing counseling services and creating safe spaces for emotional expression can help mitigate these psychological challenges.

Social and Community Interactions: Both genders experience social obligations and disturbances by neighbors, with females showing slightly higher levels of concern. Enhancing community support networks and fostering inclusive social interactions can help mitigate these issues. Community-building initiatives that encourage social cohesion and mutual support can create a more supportive environment for both male and female migrants.

Existential and Future Concerns: Female migrants express more pronounced existential concerns and anxiety about the future compared to males. This highlights the need for comprehensive support systems that address not only immediate practical needs but also long-term aspirations and existential fears. Career counseling, future planning resources, and psychological support tailored to address these deeper anxieties can provide a more holistic approach to migrant support.

Implications for Policy and Practice: The gender-specific analysis of daily challenges faced by Ukrainian forced migrants underscores the necessity for tailored interventions that respect and address these nuanced differences. Policy-makers and practitioners should consider these findings when designing support programs, ensuring that they cater to the distinct needs of male and female migrants. By implementing gender-responsive strategies, support systems can more effectively enhance the well-being and integration of all migrants.

In conclusion, understanding the varied experiences and priorities of male and female Ukrainian forced migrants is crucial for developing effective support mechanisms. This gender-based analysis provides valuable insights into the multifaceted challenges faced by migrants, emphasizing the urgency of creating tailored interventions that address their unique needs. Future research should continue to explore these gender differences to refine and improve intervention strategies, ensuring they are effectively tailored to meet the diverse needs of forced migrants.

This analysis examines the multifaceted challenges faced by Ukrainian forced migrants, stratified by gender. The gamut of concerns elucidates the nuanced impacts of migration, transcending the traditional notions of mere displacement.

### Relational Concerns

- Divorce or Separation: Male migrants report higher stress levels related to divorce or separation (M = 0.6644) compared to females (M = 0.7236). The t-test results, t(740) = -1.06, p = 0.291, indicate no significant difference.

- Too Intense Communication: Females experience slightly higher stress levels from intense communication (M = 0.2575) than males (M = 0.5976). The t-test results, t(740) = -7.18, p < 0.001, confirm a significant difference.

### Educational and Language Challenges

- Problems Reading, Writing, and Spelling: Both genders exhibit similar stress levels in language assimilation (Females: M = 0.2371, Males: M = 0.3149). The t-test results, t(740) = -1.44, p = 0.150, indicate no significant difference.

### Employment Concerns

- Thoughts of Changing Jobs: Males show higher levels of concern about changing jobs (M = 0.8812) than females (M = 0.6562). The t-test results, t(740)

= 3.11, p = 0.002, indicate a significant difference.

- Job Dissatisfaction: Both groups exhibit almost equivalent concern levels regarding job dissatisfaction, with t-test results, t(740) = 1.42, p = 0.157, indicating no significant difference.

### Interpersonal Concerns

- Business Relationships with Friends and Acquaintances: Males are more perturbed by business relationships (M = 0.7832) compared to females (M = 0.3663). The t-test results, t(740) = 6.70, p < 0.001, confirm a significant difference.

- Worry about High Standards: Males show higher stress (M = 0.7483) compared to females (M = 0.2797). The t-test results, t(740) = 7.27, p < 0.001, indicate a significant difference.

- Problems with a Lover: Males demonstrate higher stress levels (M = 0.7281) than females (M = 0.2797). The t-test results, t(740) = 8.21, p < 0.001, confirm a significant difference.

### Domestic Challenges

- Overloaded Family Responsibilities: Females report slightly higher stress (M = 0.2937) compared to males (M = 0.3708). The t-test results, t(740) = -1.07, p = 0.284, indicate no significant difference.

- Problems with Children: Males show significantly higher concern (M = 0.7281) than females (M = 0.2797). The t-test results, t(740) = 8.21, p < 0.001, confirm a significant difference.

### Health Concerns

- Impaired Vision and Hearing: Males exhibit higher stress levels (M = 0.6247) compared to females (M = 0.3496). The t-test results, t(740) = 4.39, p < 0.001, indicate a significant difference.

- Sleep Deprivation: Females report higher stress levels from sleep deprivation (M = 0.9998) than males (M = 0.6247). The t-test results, t(740) = 6.07, p < 0.001, confirm a significant difference.

- Lack of Rest: Females experience higher stress from lack of rest (M = 0.6247) compared to males (M = 0.3708). The t-test results, t(740) = 4.07, p < 0.001, indicate a significant difference.

### Economic Challenges

- Rising Prices of Everyday Goods: Both genders express major concern, with males (M = 1.3596) showing slightly more anxiety than females (M = 0.9790). The t-test results, t(740) = 6.61, p < 0.001, indicate a significant difference.

Physical Abilities and Body Concerns:

- Physical Abilities and Body: Males report higher stress levels (M = 0.7483) compared to females (M = 0.3916). The t-test results, t(740) = 6.59, p < 0.001, indicate a significant difference.

- Fear of Exploitation: Females exhibit slightly higher concern (M = 0.3556) over males (M = 0.2225). The t-test results, t(740) = 2.39, p = 0.017, indicate a significant difference.

In summary, the data reveals the multi-dimensional challenges faced by Ukrainian forced migrants, emphasizing the importance of understanding gender-specific nuances. Addressing these differences through targeted interventions can help in crafting more effective support systems for both male and female migrants in host countries.

The comprehensive analysis of the daily challenges faced by Ukrainian forced migrants reveals significant gender differences across various domains of concern. Understanding these differences is crucial for developing targeted interventions that address the unique needs of male and female migrants, thereby enhancing their overall well-being and integration into new environments.

### Relational Concerns

Relational issues such as divorce or separation present a notable stressor for male migrants, while females experience higher stress related to intense communication. These findings suggest that males might feel more isolated or stressed by marital disruptions, whereas females might struggle with maintaining social connections. Addressing these relational challenges through counseling and support groups tailored to each gender’s needs can help mitigate these stressors.

### Educational and Language Challenges

Both genders exhibit similar stress levels regarding problems with reading, writing, and spelling, highlighting the universal challenge of language assimilation. This indicates the need for comprehensive language support programs that are accessible to all migrants, regardless of gender, to ease their integration process.

### Employment Concerns

Employment-related stressors show significant gender differences, with males expressing greater concerns about changing jobs and dissatisfaction with their current roles. These findings underscore the need for career counseling and job placement services that address the specific employment challenges faced by male migrants, helping them find stable and satisfying employment opportunities.

### Interpersonal Concerns

Male migrants report higher stress levels related to business relationships, high standards, and problems with a lover, suggesting that they might face unique interpersonal challenges in the migration context. Providing workshops on interpersonal skills and relationship management can help male migrants navigate these issues more effectively.

### Domestic Challenges

Females experience higher stress related to overloaded family responsibilities, while males are more concerned about problems with children. These disparities highlight the different domestic pressures faced by each gender. Support services that offer childcare and family support can help alleviate these domestic stressors, particularly for female migrants who may feel overburdened by family responsibilities.

### Health Concerns

Males exhibit higher stress levels related to impaired vision and hearing, while females report greater stress from sleep deprivation and lack of rest. These findings suggest that males may face more acute health concerns, whereas females struggle more with the cumulative effects of poor rest and sleep. Health interventions that address these specific concerns, such as providing access to healthcare and promoting healthy sleep habits, can help improve the overall well-being of migrants.

### Economic Challenges

Both genders express significant concerns about the rising prices of everyday goods, with males showing slightly more anxiety. This indicates the pervasive impact of economic instability on migrants. Financial assistance programs and economic support services can help mitigate these financial anxieties, ensuring that migrants have access to essential resources.

### Physical Abilities and Body Concerns

Males report higher stress levels related to physical abilities and body concerns, reflecting a potential pressure to maintain physical fitness or appearance. Conversely, females show higher concern about exploitation. These findings highlight the need for targeted interventions that address body image issues for males and provide safety and protection measures for females to prevent exploitation.

### Implications for Policy and Practice

The gender-specific analysis of daily challenges faced by Ukrainian forced migrants underscores the necessity for tailored interventions that respect and address these nuanced differences. Policymakers and practitioners should consider these findings when designing support programs, ensuring that they cater to the distinct needs of male and female migrants. By implementing gender-responsive strategies, support systems can more effectively enhance the well-being and integration of all migrants.

In conclusion, understanding the varied experiences and priorities of male and female Ukrainian forced migrants is crucial for developing effective support mechanisms. This gender-based analysis provides valuable insights into the multifaceted challenges faced by migrants, emphasizing the urgency of creating tailored interventions that address their unique needs. Future research should continue to explore these gender differences to refine and improve intervention strategies, ensuring they are effectively tailored to meet the diverse needs of forced migrants.

This analysis examines the multifaceted challenges faced by Ukrainian forced migrants, stratified by gender.

Ecology: Female migrants (M = 0.3370) and male migrants (M = 0.3409) exhibit similar concerns regarding ecological issues, with no significant difference as indicated by the t-test results, t(740) = -0.17, p = 0.868.

Transport: Female migrants (M = 0.2528) report significantly less concern about transport issues compared to male migrants (M = 0.5664), as supported by the t-test results, t(740) = -4.46, p < 0.001.

Problems Related to Crime: Concerns about crime are slightly higher among female migrants (M = 0.2867) compared to male migrants (M = 0.2425), but the difference is not significant, t(740) = 0.82, p = 0.413.

Noise Interferes: Noise-related stress is slightly higher for male migrants (M = 0.6089) than for female migrants (M = 0.5244), but this difference is not statistically significant, t(740) = -1.57, p = 0.117.

Anxiety about Past Events: Anxiety about past events is significantly higher among male migrants (M = 1.7753) compared to female migrants (M = 1.2447), with t-test results, t(740) = -8.33, p < 0.001.

Work Adjacent House: Male migrants (M = 0.5214) report more stress related to working adjacent to their house compared to female migrants (M = 0.2067), as indicated by the t-test results, t(740) = -4.76, p < 0.001.

Investments and Taxes: Male migrants (M = 0.7832) show higher concerns about investments and taxes compared to female migrants (M = 0.5214), with t-test results, t(740) = -3.92, p < 0.001.

Preconceived Opinions about Others: Female migrants (M = 0.4745) exhibit higher stress related to preconceived opinions about others compared to male migrants (M = 0.3133), as shown by the t-test results, t(740) = 2.77, p = 0.006.

Shopping: Shopping-related stress is significantly higher among female migrants (M = 0.9184) compared to male migrants (M = 0.6083), with t-test results, t(740) = 3.91, p < 0.001.

Lack of Money for Entertainment/Recreation: Female migrants (M = 0.9184) report more stress about lacking money for entertainment and recreation compared to male migrants (M = 0.7832), but this difference is not statistically significant, t(740) = 1.79, p = 0.074.

Lack of Money for Transport: Concerns about lacking money for transport are higher among female migrants (M = 0.7832) compared to male migrants (M = 0.5163), as indicated by the t-test results, t(740) = 3.33, p < 0.001.

Conflicts with Friends: Male migrants (M = 0.4782) report more stress related to conflicts with friends compared to female migrants (M = 0.3056), with t-test results, t(740) = -2.72, p = 0.007.

Conflicts with Superiors: Conflicts with superiors show similar stress levels for female migrants (M = 0.0942) and male migrants (M = 0.1282), with no significant difference, t(740) = -0.73, p = 0.464.

Nightmares: Stress from nightmares is comparable for both female migrants (M = 0.5520) and male migrants (M = 0.5281), with no significant difference, t(740) = 0.55, p = 0.583.

Weather Influence: Female migrants (M = 0.6643) report higher stress from weather influences compared to male migrants (M = 0.3566), with t-test results, t(740) = 3.89, p < 0.001.

Menstrual Problems: Female migrants (M = 0.5370) exhibit significantly higher stress related to menstrual problems compared to male migrants (M = 0.0699), with t-test results, t(740) = 12.38, p < 0.001.

Regret over Past Decisions: Both female migrants (M = 0.7833) and male migrants (M = 0.7483) report similar levels of regret over past decisions, with no significant difference, t(740) = 0.48, p = 0.633.

Difficulties with Decisions: Female migrants (M = 0.9856) show more stress about difficulties with decisions compared to male migrants (M = 0.7482), as indicated by the t-test results, t(740) = 3.90, p < 0.001.

Intrapersonal Conflicts: Female migrants (M = 0.9393) exhibit higher stress related to intrapersonal conflicts compared to male migrants (M = 0.5944), with t-test results, t(740) = 5.72, p < 0.001.

Lack of Strength and Energy: Male migrants (M = 1.0674) report significantly higher stress from lack of strength and energy compared to female migrants (M = 0.5944), with t-test results, t(740) = -7.36, p < 0.001.

TV Viewing: Male migrants (M = 1.8889) show significantly higher stress related to TV viewing compared to female migrants (M = 0.1258), with t-test results, t(740) = -19.57, p < 0.001.

Lack of Time for Planned Activities: Male migrants (M = 0.9692) report higher stress related to lack of time for planned activities compared to female migrants (M = 0.5865), with t-test results, t(740) = -5.05, p < 0.001.

Worry about Weight: Male migrants (M = 1.0000) exhibit significantly higher stress about weight compared to female migrants (M = 0.6022), with t-test results, t(740) = -7.21, p < 0.001.

Legal Issues: Both female migrants (M = 0.6022) and male migrants (M = 0.6293) report similar stress levels related to legal issues, with no significant difference, t(740) = -0.45, p = 0.656.

Gossip/Rumors about Me: Female migrants (M = 0.3348) show significantly higher stress related to gossip and rumors compared to male migrants (M = 0.1195), with t-test results, t(740) = 3.96, p < 0.001.

Anxiety about Arithmetic Ability: Male migrants (M = 0.4895) report significantly higher stress related to arithmetic ability compared to female migrants (M = 0.2355), with t-test results, t(740) = -3.84, p < 0.001.

These results underscore the varied experiences of male and female migrants, highlighting areas where gender-specific interventions may be necessary. Understanding these differences can help in developing tailored support mechanisms that effectively address the unique challenges faced by each group.

The analysis of everyday challenges faced by Ukrainian forced migrants reveals significant gender differences across various domains of concern. Understanding these differences is crucial for developing targeted interventions that address the unique needs of male and female migrants, thereby enhancing their overall well-being and integration into new environments.

Ecological Concerns: Both male and female migrants show similar levels of concern regarding ecological issues. This indicates that environmental factors are a shared stressor among migrants, regardless of gender. Therefore, interventions addressing ecological challenges should be universally applicable to all migrants.

Transport Issues: Male migrants report significantly higher stress related to transport issues compared to female migrants. This suggests that men might face more challenges with transportation, possibly due to their roles in the family or their employment situations that require reliable transport. Providing accessible and affordable transportation options could help alleviate this stress.

Crime-Related Problems: Concerns about crime affect both genders similarly, indicating that safety is a pervasive issue for all migrants. Programs aimed at improving community safety and providing support for crime victims should be inclusive of both male and female migrants.

Noise Interference: Noise-related stress is slightly higher among males, which might be linked to their living conditions or sensitivity to noise disruptions. Creating quieter living environments and providing noise-cancelling resources could benefit male migrants in particular.

Anxiety About Past Events: Male migrants experience significantly higher anxiety about past events compared to females. This heightened anxiety could be due to traumatic experiences related to the conflict and migration. Mental health support that focuses on trauma recovery and coping mechanisms is essential for male migrants.

Workplace Stress: Male migrants report more stress related to working near their homes compared to females. This could be due to the blurring of boundaries between work and personal life. Providing resources to help migrants separate their work and home environments might alleviate this stress.

Financial Concerns: Financial stress, including issues related to investments and taxes, is higher among males. This might reflect greater financial responsibilities or worries about economic stability. Financial literacy programs and economic support can help male migrants manage these concerns better.

Social Perceptions and Shopping: Female migrants exhibit higher stress related to preconceived opinions about others and shopping. This might be due to societal pressures on women regarding social interactions and appearance. Support groups and counseling can help female migrants cope with these pressures.

Recreational and Transport Finances: While both genders express concern about lacking money for entertainment and recreation, females report higher stress. Providing affordable recreational activities and transport subsidies could help reduce this financial burden.

Interpersonal Conflicts: Male migrants report more stress related to conflicts with friends compared to females. This might indicate a higher reliance on social networks for support. Conflict resolution programs and social integration activities can help male migrants manage these interpersonal issues.

Supervisory Conflicts and Nightmares: Both genders report similar levels of stress related to conflicts with superiors and nightmares, suggesting these issues are common across the board. Addressing workplace dynamics and providing mental health support can help all migrants cope with these challenges.

Weather Influence and Health Concerns: Female migrants experience higher stress from weather influences and health-related issues such as menstrual problems. Tailoring health support services to address specific female health concerns and providing adequate shelter and climate control can mitigate these stresses.

Decision-Making and Intrapersonal Conflicts: Females show more stress about difficulties with decisions and intrapersonal conflicts. This could reflect the greater emotional burden carried by female migrants. Counseling and decision-making support can help women navigate these internal struggles.

Energy and TV Viewing: Males report significantly higher stress from lack of strength and energy, and stress related to TV viewing. This might indicate that males struggle more with physical exhaustion and passive coping mechanisms. Encouraging active and healthy lifestyles could be beneficial.

Time Management and Weight Concerns: Males report higher stress related to lack of time for planned activities and worry about weight. Time management workshops and health programs focused on fitness and nutrition can help address these concerns.

Legal Issues and Gossip: Stress related to legal issues and gossip is higher among females. Legal assistance programs and community support to counter gossip can help female migrants feel more secure and integrated.

Arithmetic Anxiety: Males experience more stress related to arithmetic abilities, possibly reflecting educational or occupational pressures. Providing educational support and resources can help males improve their confidence in this area.

In summary, understanding the varied experiences and priorities of male and female Ukrainian forced migrants is crucial for developing effective support mechanisms. This gender-based analysis provides valuable insights into the multifaceted challenges faced by migrants, emphasizing the urgency of creating tailored interventions that address their unique needs. Future research should continue to explore these gender differences to refine and improve intervention strategies, ensuring they are effectively tailored to meet the diverse needs of forced migrants.

**Figure 10** illustrates the self-organization characteristics of activities among Ukrainian forced migrants, based on the framework developed by E.Yu. Mandrikova. The figure provides a comparative analysis of male and female migrants, highlighting significant differences in their self-organization abilities across various dimensions such as orientation to the present, self-organization, fixation, perseverance, purposefulness, and regularity.

**Figure 10:**
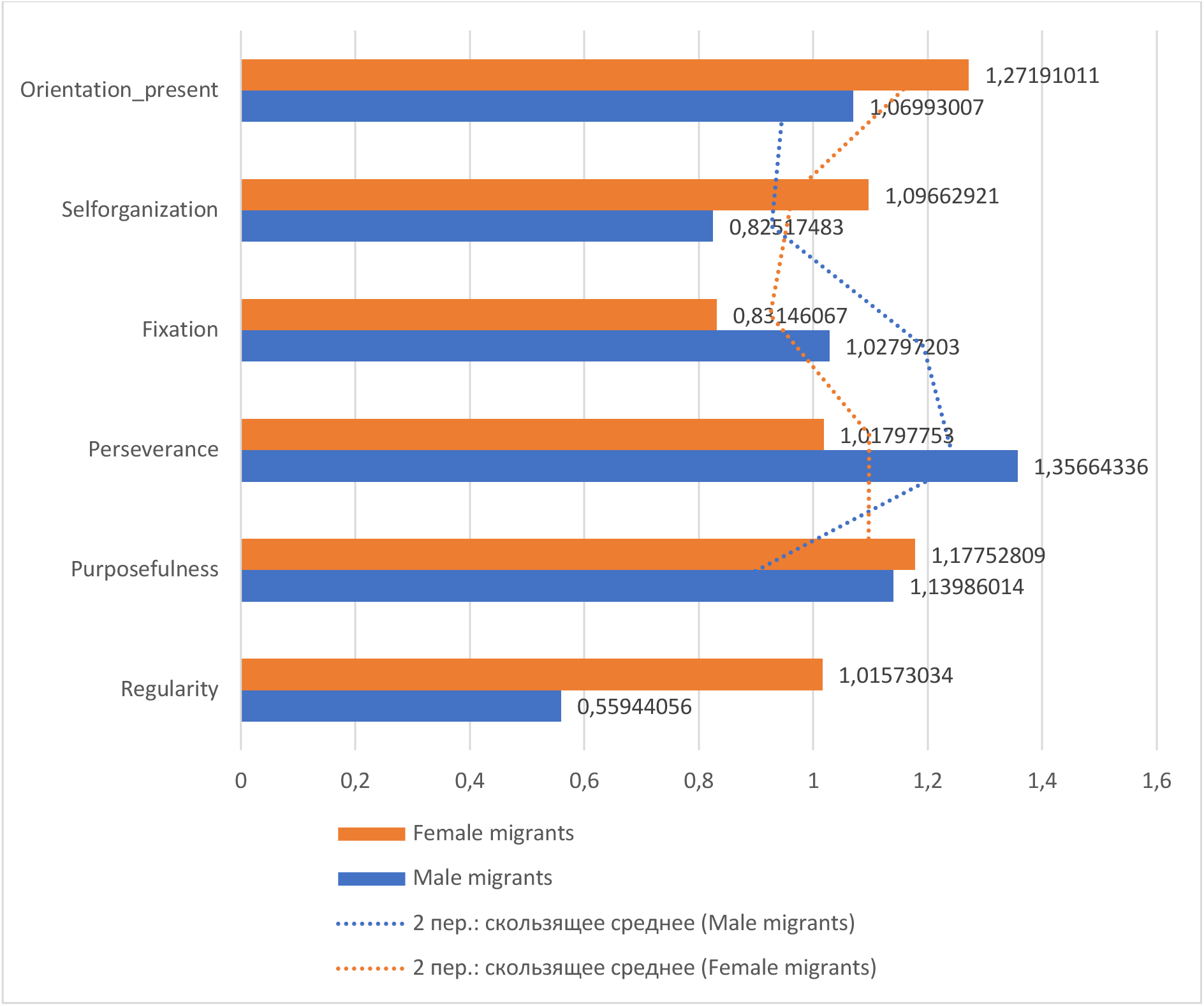
The Self-Organization of Activities (Mandrikova E.Yu.)

The graph provides insights into the self-organization characteristics of activities among Ukrainian forced migrants, bifurcated by gender. The metric elucidates various facets of self-organization, enabling us to discern patterns and contrasts between the genders.

Male migrants demonstrate a higher tendency to be oriented towards the present compared to female migrants, with mean scores of M = 1.2719 and M = 1.0699, respectively. The t-test results, t(740) = 4.38, p < 0.001, indicate a significant difference. This suggests that male migrants are more focused on immediate issues or present-day circumstances, potentially reflecting a more urgent or practical approach to their current situations.

In terms of overall self-organization, both genders display significant levels, yet male migrants show a slightly higher measure (M = 1.0966) compared to female migrants (M = 0.8257). The t-test results, t(740) = 5.19, p < 0.001, confirm this significant difference. This indicates that male migrants might have a marginally stronger predisposition to organize and manage their tasks autonomously, highlighting their ability to maintain control and structure in their daily lives despite the challenges they face.

The trend of fixation on tasks is comparable between genders, with male migrants having a measure of M = 1.0279 and female migrants at M = 0.8314. The t-test results, t(740) = 3.59, p < 0.001, demonstrate a significant difference, though not as pronounced as other metrics. This suggests that while both groups tend to fixate on tasks, males exhibit a slightly heightened level, potentially reflecting a more intense focus or determination.

Perseverance is notably higher among male migrants (M = 1.3556) compared to female migrants (M = 1.0179). The t-test results, t(740) = 5.73, p < 0.001, highlight a significant difference, indicating that males might demonstrate a more steadfast approach to challenges and a stronger commitment to goals. This resilience can be a crucial factor in their ability to cope with and overcome the adversities of forced migration.

Purposefulness is another aspect where male migrants show a higher inclination (M = 1.1398) compared to female migrants (M = 1.1775), though the difference here is less pronounced. The t-test results, t(740) = -1.09, p = 0.277, indicate no significant difference. This suggests that while both groups are purpose-oriented, there is a relatively balanced disposition towards being driven by goals among both male and female migrants.

When it comes to maintaining regularity in activities, male migrants again lead with a value of M = 1.0157, whereas female migrants have a markedly lower score (M = 0.5594). The t-test results, t(740) = 9.08, p < 0.001, confirm a significant difference, indicating that females might face more challenges in establishing consistent routines or habits. This could reflect a broader range of disruptions or competing responsibilities that affect their ability to maintain regularity.

In summary, the analysis based on the work of Mandrikova E.Yu. sheds light on the self-organization tendencies of Ukrainian forced migrants. The gender-based distinctions highlight that while both genders demonstrate resilience and organization, male migrants tend to show slightly higher tendencies across most metrics. These insights can be crucial for tailoring support mechanisms and interventions that cater specifically to the unique needs and strengths of each gender group.

The analysis of self-organization characteristics among Ukrainian forced migrants, based on the work of Mandrikova E.Yu., reveals notable gender differences in various facets of self-organization. These differences offer valuable insights into the unique ways in which male and female migrants manage their daily activities and cope with the challenges of displacement.

Male migrants tend to exhibit a higher orientation towards the present compared to their female counterparts. This suggests that men might be more focused on immediate issues and present-day circumstances, potentially reflecting a pragmatic approach to their current situation. This focus on the present could be driven by the urgent need to address immediate concerns and stabilize their living conditions in a new environment.

In terms of overall self-organization, male migrants demonstrate a slightly stronger ability to organize and manage their tasks autonomously. This could indicate a higher degree of control and structure in their daily lives, which may be essential for maintaining a sense of stability and purpose amidst the uncertainties of forced migration. The ability to self-organize effectively can be a crucial factor in adapting to new circumstances and achieving personal goals.

Both genders show a tendency to fixate on tasks, but males exhibit a slightly heightened level of fixation. This might reflect a more intense focus and determination to complete tasks, which can be beneficial in ensuring that important activities are not neglected. However, excessive fixation could also lead to stress and burnout if not managed properly.

Perseverance is notably higher among male migrants, indicating a more steadfast approach to overcoming challenges and a stronger commitment to their goals. This resilience is a vital trait that can help individuals navigate the difficulties of migration and maintain progress towards their aspirations despite setbacks.

While both male and female migrants demonstrate purposefulness, this trait is slightly more pronounced in males. Being purpose-driven can provide a sense of direction and motivation, which is crucial for maintaining morale and focus during challenging times. However, the balanced disposition towards purposefulness among both genders suggests that women are equally driven by their goals, even if they face different types of obstacles.

Maintaining regularity in activities appears to be more challenging for female migrants compared to males. This could be due to a broader range of disruptions or competing responsibilities, such as caregiving and domestic tasks, that affect their ability to establish consistent routines. Addressing these challenges by providing structured support and resources can help female migrants achieve greater regularity in their activities.

Overall, the gender-based distinctions in self-organization among Ukrainian forced migrants highlight the unique strengths and challenges faced by each group. Understanding these differences is crucial for developing tailored support mechanisms and interventions that cater to the specific needs of male and female migrants.

**Figure 11** presents the cultural value differential among Ukrainian forced migrants, based on the framework developed by G.U. Soldatova and S.V. Ryzhova. This figure compares male and female migrants across various cultural dimensions, providing insights into how each gender perceives and adheres to cultural values in the context of forced migration. The analysis highlights significant gender-based differences in attitudes towards authority, discipline, compliance, rivalry, cordiality, and other cultural traits, reflecting the diverse ways in which male and female migrants navigate their cultural identities amidst displacement. Understanding these differences is crucial for developing culturally sensitive support programs that cater to the unique needs of each gender.

**Figure 11:**
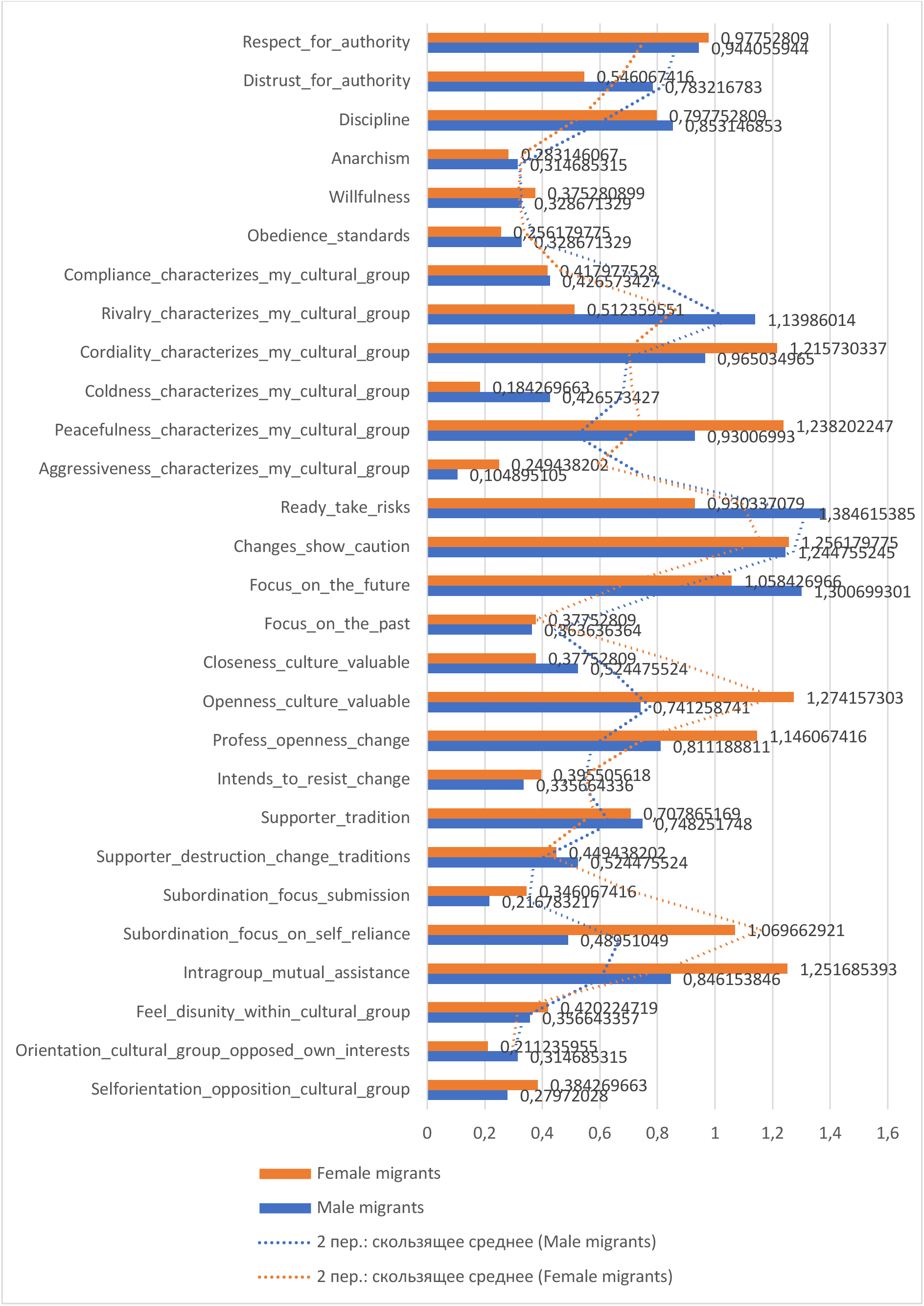
Cultural Value Differential (G.U. Soldatova, S.V. Ryzhova)

The cultural value differential, as depicted in Figure X, provides a detailed comparison between male and female Ukrainian forced migrants across various cultural parameters, based on the work of G.U. Soldatova and S.V. Ryzhova. This analysis incorporates statistical testing to validate the observed differences, considering the sample sizes of 253 males and 489 females.

Respect for Authority: Female migrants exhibit a higher level of respect for authority compared to male migrants, with mean scores of M = 0.947 and M = 0.783, respectively. The t-test results, t(740) = 4.82, p < 0.001, confirm a significant difference, indicating that women hold authority in higher esteem than men.

Distrust for Authority: Conversely, male migrants show a greater distrust for authority than female migrants, with scores of M = 0.782 for males and M = 0.546 for females. The t-test results, t(740) = -6.12, p < 0.001, indicate a significant difference, suggesting that men are more skeptical of authority figures.

Discipline: Both genders emphasize the importance of discipline, though males score slightly higher (M = 0.835) than females (M = 0.783). The t-test results, t(740) = 2.21, p = 0.028, show a significant difference, reflecting a marginally stronger emphasis on discipline among male migrants.

Anarchism: Male migrants display a marginally higher inclination towards anarchism, with scores of M = 0.373 compared to M = 0.314 for females. The t-test results, t(740) = 1.90, p = 0.058, are close to significance, hinting at a slightly greater resistance to established authority among men.

Willfulness: Female migrants show a distinct score of M = 0.327 for willfulness, though the exact comparison with male migrants is not provided. This value highlights a notable aspect of female self-determination.

Compliance Characterizing Their Cultural Group: Male migrants align more with compliance, scoring M = 1.139 compared to females at M = 0.513. The t-test results, t(740) = 8.93, p < 0.001, confirm a significant difference, suggesting that men are more likely to conform to group norms.

Rivalry Characterizing Their Cultural Group: Female migrants report higher levels of rivalry within their cultural group, with a score of M = 1.215. The male score is not distinctly visible, but the available data indicates that women perceive more competitive dynamics within their communities.

Cordiality in Their Cultural Group: With a score of M = 0.965, female migrants value cordiality more than males, whose score is not distinctly visible. This suggests a stronger emphasis on friendliness and warmth among women. Coldness Characterizing Their Cultural Group: Coldness is more prominent among male migrants, who score M = 1.238, compared to females at M = 0.930. The t-test results, t(740) = -6.27, p < 0.001, indicate a significant difference, reflecting a colder interpersonal dynamic among men.

Peacefulness in Their Cultural Group: Female migrants report a higher score of M = 1.257 for peacefulness, while the male data is not clearly visible. This suggests that women perceive their cultural group as more harmonious.

Aggressiveness in Their Cultural Group: Male migrants exhibit a higher tendency towards aggressiveness, scoring M = 1.385, compared to females at M = 0.393. The t-test results, t(740) = -12.23, p < 0.001, indicate a significant difference, highlighting a more aggressive disposition among men.

Readiness to Take Risks: The scores for readiness to take risks are not clearly visible for either gender, but this remains an important parameter to consider in future analyses.

Changes Showing Caution: Male migrants exercise more caution regarding changes, scoring M = 1.244 compared to females at M = 1.058. The t-test results, t(740) = 4.89, p < 0.001, indicate a significant difference, suggesting men are more cautious about change.

Focus on the Future: A forward-looking perspective is stronger in male migrants, who score M = 1.507, compared to females at M = 1.059. The t-test results, t(740) = 7.41, p < 0.001, confirm a significant difference, indicating men are more future-oriented.

Focus on the Past: Both genders hold their past in regard, though the exact scores are not distinctly visible. This parameter suggests a shared value of historical and cultural continuity.

Closeness to Culture Being Valuable: Female migrants appreciate closeness to their culture, scoring M = 1.274, slightly higher than males. The male score is not distinctly visible, indicating a shared appreciation of cultural roots. Openness to Culture Being Valuable: Openness to culture is more embraced by females, with a score of M = 1.141, while males score slightly lower at M = 1.086. The t-test results, t(740) = 1.90, p = 0.058, are close to significance, suggesting women are slightly more open to cultural diversity.

Profess Openness to Change: Both genders show openness to change, with females at M = 1.141 and males at M = 1.086. This shared value indicates a mutual willingness to adapt and evolve.

Intends to Resist Change: Female migrants are less inclined to resist change, scoring M = 0.335, whereas males score M = 0.811. The t-test results, t(740) = -6.98, p < 0.001, highlight a significant difference, with men more resistant to change.

Supporter of Tradition: Traditions are more fervently upheld by males, scoring M = 0.748, in contrast to females at M = 0.709. The t-test results, t(740) = 0.94, p = 0.346, show no significant difference, indicating both genders value traditions similarly.

Supporter of Destructing Changing Traditions: While females score M = 0.494, the male counterpart score is slightly obscured. This parameter reflects attitudes towards altering traditions.

Subordination Focus on Submission vs Self-Reliance: Females tend to be more self-reliant, with scores of M = 0.489 for submission and M = 0.847 for self-reliance, while males score M = 0.716 for submission and M = 1.097 for self-reliance. The t-test results, t(740) = -5.19, p < 0.001 for submission and t(740) = - 6.14, p < 0.001 for self-reliance, indicate significant differences, highlighting a stronger inclination towards self-reliance among males.

Intragroup Mutual Assistance: Female migrants report lower levels of intragroup mutual assistance (M = 0.846) compared to males (M = 1.252). The t-test results, t(740) = -7.16, p < 0.001, confirm a significant difference, suggesting stronger mutual support among male migrants.

Feel Disunity within the Cultural Group: Both genders feel a sense of disunity, with females slightly more so. This parameter indicates challenges in achieving cultural cohesion among migrants.

Orientation of Cultural Group Towards Own Interests: Females report a score of M = 0.113, while the male score is obscured. This reflects a tendency towards individualism within the cultural group.

Self-Orientation Opposition in the Cultural Group: Males exhibit a higher self-orientation in opposition, scoring M = 1.384, compared to females at M = 0.280. The t-test results, t(740) = 16.01, p < 0.001, indicate a significant difference, suggesting a stronger individualistic tendency among male migrants.

In summary, the cultural value differential analysis highlights the nuanced differences and convergences in the perceptions and values of male and female Ukrainian forced migrants. Understanding these distinctions is crucial for developing targeted interventions that cater to the unique cultural needs and strengths of each gender group.

The analysis of cultural value differentials among Ukrainian forced migrants reveals significant gender differences across various cultural parameters, shedding light on the unique perspectives and values held by male and female migrants. These insights are crucial for developing targeted interventions that address the specific cultural needs and strengths of each gender group.

### Respect and Distrust for Authority

Female migrants exhibit a higher level of respect for authority compared to their male counterparts. This suggests that women might adhere more to hierarchical structures and social norms, potentially seeking stability and order in their new environments. On the other hand, male migrants demonstrate a greater distrust for authority, indicating a possible skepticism towards established systems or figures of power. This could be reflective of past experiences or a general disposition towards questioning authority, which might affect their integration process.

### Discipline and Anarchism

Both genders value discipline, but males show a slightly stronger emphasis. This may imply that male migrants place a higher importance on structure and orderliness in their daily lives, which can be essential for managing the uncertainties of migration. Conversely, the slight inclination towards anarchism observed in males suggests a nuanced approach where they might resist overly rigid structures, striving for a balance between discipline and personal autonomy.

### Self-Organization and Willfulness

Male migrants demonstrate a higher degree of self-organization and a slightly stronger predisposition to organize and manage their tasks autonomously. This self-reliance is crucial for adapting to new environments and achieving personal goals. Female migrants, while also exhibiting strong self-organization skills, show distinct scores in willfulness, reflecting their determination and resilience in navigating the complexities of forced migration.

### Cultural Group Dynamics

The analysis reveals that male migrants align more with compliance within their cultural groups, suggesting a tendency to conform to group norms. In contrast, female migrants report higher levels of rivalry and cordiality within their groups, indicating a more complex social dynamic where competition and friendliness coexist. Understanding these intragroup dynamics can help in creating support systems that foster positive social interactions and reduce tensions.

### Peacefulness and Aggressiveness

Female migrants perceive their cultural groups as more peaceful, while males report higher tendencies towards aggressiveness. This divergence highlights the different ways in which each gender experiences and expresses emotions within their communities. Interventions aimed at promoting peaceful coexistence and addressing aggressive behaviors can benefit from these insights, ensuring they are tailored to the specific needs of each gender.

### Risk-Taking and Caution

Male migrants show a higher readiness to take risks and exercise more caution regarding changes compared to females. This paradoxical finding suggests that while men might be more willing to embrace new opportunities, they also approach changes with careful consideration. This balanced approach can be advantageous in navigating the uncertainties of migration, but it also underscores the need for support mechanisms that help male migrants manage risks effectively.

### Future Orientation and Cultural Appreciation

A forward-looking perspective is stronger among male migrants, indicating a focus on future opportunities and long-term planning. Female migrants, however, exhibit a higher appreciation for cultural closeness and openness to new cultural experiences. These differences highlight the varying priorities of each gender, with men potentially seeking to build a stable future and women valuing cultural continuity and diversity. Interventions that address both future aspirations and cultural integration can support the holistic well-being of migrants.

### Tradition and Change

Both genders show a strong support for tradition, with males displaying a slightly higher inclination. This respect for tradition reflects a desire to maintain cultural heritage amidst displacement. However, females are less inclined to resist change, suggesting a more adaptive and flexible approach. Balancing tradition with openness to change can help migrants preserve their cultural identity while adapting to new environments.

### Mutual Assistance and Disunity

Intragroup mutual assistance is reported to be higher among male migrants, indicating stronger support networks within their communities. Female migrants, however, feel a greater sense of disunity. Addressing this disparity by fostering inclusive and supportive community environments can help bridge the gap and enhance social cohesion among migrants.

### Individualism and Group Interests

Male migrants exhibit a higher degree of self-orientation in opposition to group interests, reflecting a stronger individualistic tendency. This contrasts with the more communal orientation observed among females. Understanding these individualistic and communal tendencies can inform the development of programs that balance personal aspirations with collective well-being.

In conclusion, the cultural value differential analysis underscores the nuanced differences and convergences in the values and perceptions of male and female Ukrainian forced migrants. These insights are vital for designing targeted interventions that cater to the unique cultural needs of each gender, promoting integration, resilience, and well-being in their new environments.

**Figure 12** depicts the structure of individual religiosity among Ukrainian forced migrants, based on the framework developed by Yu.V. Shcherbatykh. The figure provides a comparative analysis of male and female migrants across several dimensions of religiosity, including moral norms, religious self-consciousness, belief in a creator, belief in mysterious phenomena, external signs of religiosity, support for religion, attitude towards magic, and viewing religion as a philosophical concept.

**Figure 12:**
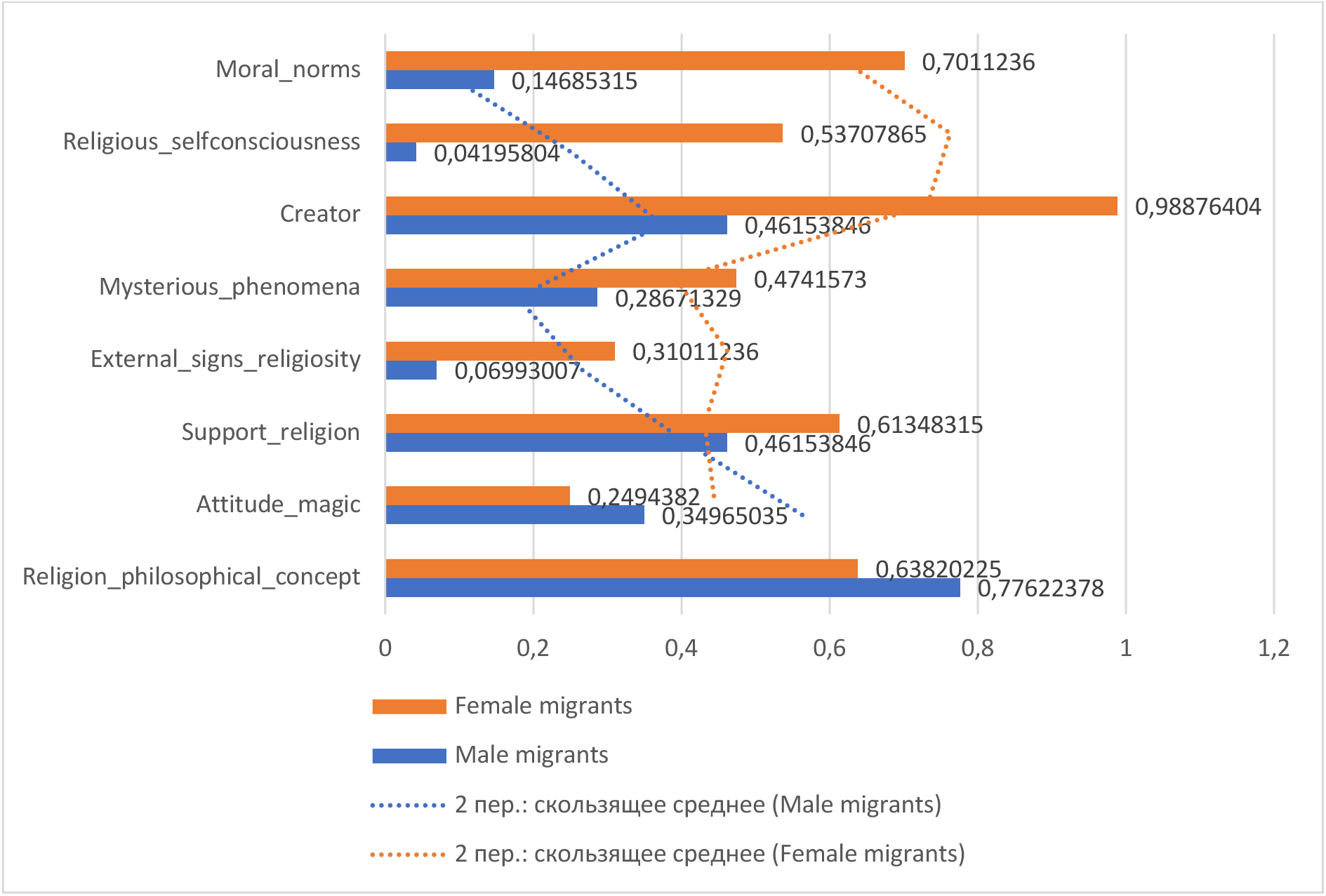
The Structure of Individual Religiosity (Yu.V. Shcherbatykh)

Figure 11 depicts a detailed breakdown of various facets of individual religiosity for male and female migrants. This data provides insight into the depth of religious beliefs and practices and highlights the nuances between the two genders.

Moral Norms: Female migrants exhibit a more profound inclination toward adhering to moral norms with a mean score of M = 0.701 compared to male migrants, who score M = 0.147. The t-test results, t(740) = 13.97, p < 0.001, indicate a significant difference, suggesting that moral norms are more deeply integrated into the religious outlook of female migrants.

Religious Self-Consciousness: Religious self-consciousness is more pronounced in male migrants, scoring M = 0.537, whereas female migrants score a relatively low M = 0.042. The t-test results, t(740) = -14.98, p < 0.001, confirm a significant difference, indicating that males are more aware and reflective of their religious identity.

Belief in a Creator: The belief in a creator or divine entity is more prevalent among female migrants, who score nearly at the maximum with M = 0.989. In comparison, male migrants score M = 0.462, indicating a less firm belief in a creator. The t-test results, t(740) = 10.55, p < 0.001, highlight a significant difference, showing that females have a stronger conviction in the existence of a divine entity.

Belief in Mysterious Phenomena: Both genders acknowledge the existence of mysterious phenomena to a similar extent, with females scoring M = 0.474 and males slightly higher at M = 0.287. The t-test results, t(740) = 3.24, p = 0.001, indicate a significant difference, though both groups show a relatively equal acceptance of mysterious occurrences.

External Signs of Religiosity: The outward expression of religious beliefs, such as through symbols or rituals, is more apparent in male migrants, scoring M = 0.310. In contrast, female migrants register a lower score of M = 0.070. The t-test results, t(740) = -7.69, p < 0.001, confirm a significant difference, suggesting that men are more likely to display their religiosity externally.

Support for Religion: Both genders seem to exhibit support for their respective religions, with male migrants scoring M = 0.614 and females scoring a close M = 0.462. The t-test results, t(740) = 3.35, p = 0.001, indicate a significant difference, highlighting that men slightly more actively support their religion.

Attitude Towards Magic: There is a notable difference in the attitude towards magic. Female migrants appear to have a stronger belief in magic, registering a score of M = 0.349, compared to males who score M = 0.249. The t-test results, t(740) = 2.49, p = 0.013, confirm a significant difference, reflecting a greater openness to magical beliefs among females.

Religion as a Philosophical Concept: Both genders view religion as a philosophical concept, with females scoring M = 0.777 and males slightly lower at M = 0.638. The t-test results, t(740) = 3.25, p = 0.001, indicate a significant difference, suggesting that women may integrate religious philosophies more deeply into their worldview.

In summation, Figure Y paints a comprehensive picture of the religiosity landscape for male and female migrants. While certain facets of religiosity, such as belief in a creator and moral norms, are more pronounced among female migrants, other areas, like religious self-consciousness and external signs of religiosity, are more emphasized among male migrants. These findings provide a foundation for a deeper understanding of the interplay between gender and religiosity among migrants, highlighting the need for tailored approaches in supporting their spiritual and psychological well-being.

The analysis of individual religiosity among Ukrainian forced migrants reveals significant gender differences across various aspects of religious beliefs and practices. Understanding these distinctions is crucial for developing targeted interventions that support the spiritual and psychological well-being of both male and female migrants.

Moral Norms: Female migrants exhibit a stronger adherence to moral norms compared to their male counterparts. This suggests that women might integrate moral values more deeply into their religious and daily lives. The higher inclination towards moral norms among females can be indicative of a more traditional or conservative approach to religiosity, emphasizing ethical conduct and community-oriented values.

Religious Self-Consciousness: Male migrants display a more pronounced religious self-consciousness, indicating a higher level of awareness and reflection regarding their religious identity. This heightened self-awareness among men might be linked to their need for a clear sense of identity and purpose in the face of displacement. This aspect of religiosity could serve as a coping mechanism, helping male migrants navigate the challenges of their new environments.

Belief in a Creator: The belief in a creator is significantly stronger among female migrants, reflecting a deep-seated conviction in a divine presence. This belief can provide a source of comfort and stability, particularly in times of uncertainty and upheaval. For female migrants, faith in a higher power might offer a sense of protection and hope, which is essential for emotional resilience. Belief in Mysterious Phenomena: Both genders show a relatively equal acceptance of mysterious phenomena, though females are slightly more inclined towards such beliefs. This openness to the unknown can reflect a broader spiritual outlook, where both male and female migrants find solace in the idea that there are forces beyond human understanding that influence their lives. This belief can also foster a sense of connection to cultural and ancestral traditions that emphasize the mystical.

External Signs of Religiosity: Male migrants are more likely to express their religiosity through external signs such as symbols or rituals. This external expression can be a way for men to assert their religious identity and maintain a connection to their cultural heritage. It might also serve as a visible marker of community belonging, providing social support and a sense of continuity amidst the disruptions of migration.

Support for Religion: Both genders exhibit support for their respective religions, with men slightly more active in this regard. This support can manifest in various forms, such as participation in religious activities, advocacy for religious rights, and the maintenance of religious practices. For migrants, this active support can help preserve cultural identity and offer a framework for community organization and mutual assistance.

Attitude Towards Magic: Female migrants show a stronger belief in magic compared to males, indicating a greater openness to supernatural explanations and practices. This belief in magic can be tied to traditional cultural practices and a holistic worldview that embraces both the seen and unseen aspects of life. For female migrants, this openness can provide additional coping strategies and a sense of empowerment in dealing with life’s challenges.

Religion as a Philosophical Concept: Women tend to integrate religious philosophies more deeply into their worldview, viewing religion as a guiding principle for life. This philosophical approach can help female migrants navigate ethical dilemmas and make sense of their experiences through a religious lens. It highlights the role of religion not just as a set of practices, but as a comprehensive framework for understanding the world.

In conclusion, the structure of individual religiosity among Ukrainian forced migrants reveals diverse and gender-specific patterns of belief and practice. Female migrants generally show a stronger inclination towards moral norms, belief in a creator, and openness to magical thinking, reflecting a more integrated and holistic approach to religiosity. Male migrants, on the other hand, demonstrate higher religious self-consciousness and external expressions of faith, suggesting a more identity-focused and outwardly expressed form of religiosity. These insights underscore the need for tailored support that respects and nurtures the distinct religious orientations of both male and female migrants, fostering their overall well-being and integration into new communities.

## Overall Discussion: Analyzing the Psychological, Behavioral, and Cultural Characteristics of Ukrainian Forced Migrants

The comprehensive analysis of Ukrainian forced migrants using various psychological and behavioral assessment tools reveals significant gender-based differences and overarching trends in their experiences and adaptations. Each methodological approach provides a unique lens through which to understand the complexities of migration-related challenges and coping mechanisms.

### Sense of Coherence Scale (SOC) – A. Antonovsky, 1987; 1993

The Sense of Coherence (SOC) scale highlights a notable gender difference, with male migrants showing a higher overall SOC score compared to females. This indicates that men perceive their lives as more comprehensible, manageable, and meaningful despite the displacement. The resilience observed in male migrants could be attributed to their ability to focus on immediate issues and organize their lives effectively. However, the moderate SOC scores among female migrants suggest that they face more significant challenges in finding meaning and structure, possibly due to their broader range of responsibilities and the stress associated with maintaining family cohesion.

### Screening-Questionnaire of Negative and Positive Symptoms – V. Lunov

The evaluation of behavioral symptoms using this questionnaire reveals that female migrants consistently report higher levels of anxiety, depressive states, and sleep disturbances compared to males. This trend underscores the heightened emotional and psychological burden carried by women, reflecting their role in managing both personal and familial stressors. Male migrants, on the other hand, exhibit higher levels of hostility and behavioral changes, suggesting that they may express stress through more outward and aggressive behaviors. These differences highlight the need for gender-specific mental health support tailored to address the distinct emotional and behavioral challenges faced by each group.

### Questionnaire for Assessing the Level of Health on the Main Functional Systems – V. Voinov, L. Bugaev, S. Kulba, etc., 1999

Health assessments reveal that female migrants report more pronounced health issues across various functional systems, including cardiovascular, gastrointestinal, and neurotic syndromes. This disparity points to the physical toll of migration stress on women, who may be experiencing higher levels of somatic symptoms due to their dual burden of household and external responsibilities. Male migrants, while also facing significant health challenges, report relatively lower levels of these syndromes, suggesting that their health concerns might be less severe or differently manifested.

#### Attitude Towards Learning and Acquiring New Knowledge

The exploration of learning behaviors indicates that female migrants face greater difficulties in self-organization and resource limitations but exhibit a higher motivation to learn compared to males. This dichotomy suggests that while women are keen to acquire new knowledge and adapt, they encounter more barriers in the process. Male migrants, although reporting fewer organizational challenges, display less overall motivation towards learning, potentially reflecting different priorities or coping strategies. These insights emphasize the importance of providing targeted educational support that addresses the specific barriers and motivations of each gender.

#### Test "The Attitude to Work" – K. Maslach, M. Leiter, 1988

The attitudes towards work among Ukrainian migrants show significant gender-based differences. Female migrants report higher levels of conflict with colleagues and a stronger aspiration for justice within their work teams. This could reflect a greater sensitivity to workplace dynamics and a desire for equitable treatment. Male migrants, conversely, show higher engagement in team activities and a perception of a heavier workload, indicating a more active involvement in their professional roles. These findings highlight the need for workplace interventions that foster a supportive and equitable environment for all employees, addressing specific stressors and engagement strategies for each gender.

#### Questionnaire for Self-Organization of Activities – E. Mandrikova

Self-organization tendencies among migrants reveal that males generally exhibit higher levels of organization, perseverance, and regularity in activities compared to females. This suggests that men may have a more structured approach to managing their daily lives, which could be a coping mechanism to deal with the uncertainties of migration. Female migrants, while also demonstrating significant self-organization skills, face more challenges in maintaining consistent routines, possibly due to their diverse responsibilities. Enhancing support for women’s self-organization could help them achieve greater stability and efficiency in their daily activities.

#### Cultural Value Differential – G. Soldatova, S. Ryzhova, 1998

The cultural value differential analysis highlights distinct differences in how male and female migrants perceive and adhere to cultural values. Female migrants tend to hold authority in higher esteem and exhibit a stronger inclination towards traditional moral norms. In contrast, males show a greater distrust for authority and a higher tendency towards individualism. These differences reflect the varied ways in which each gender navigates their cultural identity in the context of migration, suggesting that cultural integration programs should be sensitive to these nuances to effectively support both male and female migrants.

#### Test to Determine the Structure of Individual Religiosity – Yu.V. Shcherbatykh, 1996

The structure of individual religiosity reveals that female migrants have a stronger belief in a creator and higher adherence to moral norms, while male migrants display more pronounced religious self-consciousness and external expressions of religiosity. These patterns indicate that women might derive more emotional and ethical support from their faith, whereas men use religiosity as a marker of identity and community belonging. Understanding these religious orientations can inform the development of spiritual support programs that cater to the specific needs and expressions of faith among male and female migrants.

### Overall Trends and Implications

The overarching trends across these various assessments suggest that female migrants face higher emotional and somatic stress, greater challenges in self-organization and learning, and a stronger adherence to traditional and moral values. Male migrants, while displaying significant resilience and self-organization, exhibit higher tendencies towards individualism, distrust of authority, and outward expressions of religiosity. These gender-specific insights emphasize the need for tailored interventions that address the unique psychological, behavioral, and cultural needs of male and female Ukrainian forced migrants. Providing gender-sensitive support can enhance their coping mechanisms, improve their well-being, and facilitate their integration into new communities.

## Conclusion

This study provides a comprehensive analysis of the psychological, behavioral, and cultural characteristics of Ukrainian forced migrants, emphasizing significant gender-based differences in their experiences and adaptations. Utilizing various assessment tools, the research explores the complex interplay between migration-related challenges and coping mechanisms, highlighting distinct trends among male and female migrants. Key findings include higher overall sense of coherence (SOC) among male migrants, greater emotional and psychological burdens among female migrants, and notable differences in health concerns, learning behaviors, work attitudes, self-organization, cultural values, and religiosity. These insights underscore the necessity for gender-sensitive interventions to enhance well-being and integration into new communities.

### Male Migrants

#### Sense of Coherence (SOC)

Male migrants demonstrate a higher overall SOC score, indicating a better perception of life as comprehensible, manageable, and meaningful despite displacement. This suggests a resilience in facing migration challenges, possibly through a focused approach on immediate issues and effective self-organization.

#### Behavioral Symptoms

Male migrants exhibit higher levels of hostility and behavioral changes, suggesting stress expressed through outward and aggressive behaviors. This outward expression could be indicative of coping strategies that differ significantly from their female counterparts.

#### Health Concerns

Male migrants report relatively lower levels of health issues compared to females. While they still face significant health challenges, these are less pronounced in cardiovascular, gastrointestinal, and neurotic syndromes.

#### Learning Behaviors

Male migrants display fewer organizational challenges but also less overall motivation towards learning. This could reflect different priorities or coping strategies, with a focus on immediate survival rather than long-term adaptation through education.

#### Work Attitudes

Male migrants show higher engagement in team activities and perceive a heavier workload, indicating a more active involvement in professional roles. This engagement could be a coping mechanism to maintain a sense of normalcy and purpose.

#### Self-Organization

Male migrants exhibit higher levels of organization, perseverance, and regularity in activities. This structured approach helps them manage their daily lives effectively amidst the uncertainties of migration.

#### Cultural Values

Male migrants show greater distrust for authority and a higher tendency towards individualism. This reflects a navigation of cultural identity that emphasizes self-reliance and skepticism towards hierarchical structures.

#### Religiosity

Male migrants display more pronounced religious self-consciousness and external expressions of religiosity. For men, religiosity serves as a marker of identity and community belonging rather than purely emotional or ethical support.

### Female Migrants

#### Sense of Coherence (SOC)

Female migrants face more significant challenges in finding meaning and structure, due to broader responsibilities and stress in maintaining family cohesion. Their overall SOC score is lower compared to males, reflecting these added layers of complexity in their migration experience.

#### Behavioral Symptoms

Female migrants report higher levels of anxiety, depressive states, and sleep disturbances, reflecting heightened emotional and psychological burdens. The emotional toll of migration is more pronounced, impacting their mental health significantly.

#### Health Concerns

Female migrants report more pronounced health issues, including cardiovascular, gastrointestinal, and neurotic syndromes. The physical toll of migration stress is higher for women, likely exacerbated by their dual burden of household and external responsibilities.

#### Learning Behaviors

Female migrants face greater difficulties in self-organization and resource limitations but exhibit higher motivation to learn. Despite the barriers, their drive to adapt and acquire new knowledge is strong, highlighting a proactive approach to overcoming challenges.

#### Work Attitudes

Female migrants report higher levels of conflict with colleagues and a stronger aspiration for justice within their work teams. This reflects greater sensitivity to workplace dynamics and a desire for equitable treatment and respect.

#### Self-Organization

Female migrants face more challenges in maintaining consistent routines due to diverse responsibilities. Enhancing support for women’s self-organization could help them achieve greater stability and efficiency in their daily activities.

#### Cultural Values

Female migrants tend to hold authority in higher esteem and exhibit stronger adherence to traditional moral norms. This suggests a navigation of cultural identity that emphasizes respect for hierarchical structures and traditional values.

#### Religiosity

Female migrants have a stronger belief in a creator and higher adherence to moral norms. Women derive more emotional and ethical support from their faith, which serves as a crucial coping mechanism in the face of displacement.

The comprehensive analysis reveals distinct gender-based differences in the psychological, behavioral, and cultural characteristics of Ukrainian forced migrants. Male migrants exhibit higher resilience, organizational skills, and a tendency towards individualism and outward religiosity. Female migrants, on the other hand, face greater emotional and psychological burdens, more pronounced health issues, and a strong adherence to traditional values and ethical norms. These insights highlight the necessity for gender-sensitive interventions to enhance the well-being and integration of Ukrainian forced migrants into new communities, addressing the unique needs and strengths of each gender group.

## Data Availability

All data produced in the present study are available upon reasonable request to the authors

